# The germinal center B cell response to pneumococcal conjugate vaccines is antigenically restricted

**DOI:** 10.64898/2026.05.12.26353001

**Authors:** D.W. de Vos, M. Johnson, D. Hoving, G.E. Loe-Sack-Sioe, C. Kienhuis, E.L. van Persijn van Meerten, D. Goldblatt, L.G. Visser, A.H.E. Roukens, S.P. Jochems

## Abstract

Despite the availability of effective vaccines, pneumococcal disease remains a major global health concern. Pneumococcal vaccines are multivalent vaccines that have progressively increased in valency, a change associated with lower antibody titers to individual polysaccharide antigens. Whether increasing vaccine valency influences B cell responses through antigenic competition remains incompletely understood. Here, we studied pneumococcal polysaccharide-specific B cell responses in peripheral blood and lymph nodes of healthy adults following vaccination with the 13-valent pneumococcal conjugate vaccine. Antigen-specific memory B cells in peripheral blood expanded 2 weeks post-vaccination, whereas germinal center formation was delayed and peaked after 4 weeks. Notably, germinal center B cell responses were dominated by a limited number of specificities, in contrast to the more evenly distributed expansion observed in peripheral blood. Together, these data highlight the importance of extrafollicular responses in adult anti-pneumococcal polysaccharide immunity and provide evidence for antigenic competition during lymph node germinal center formation, which may have important implications in the context of multivalent vaccines.

**Summary:** Using lymph node fine needle aspiration, the study characterizes germinal center responses to multivalent pneumococcal conjugate vaccines. Antigen-specific peripheral blood B cells expand before germinal center formation, and germinal center B cells display a restricted serotype-specific response.

## Introduction

*Streptococcus pneumoniae* is a pathogen causing a substantial burden of disease globally. Over 100 *S. pneumoniae* serotypes have been described to date (1). *S. pneumoniae* commonly colonizes the upper respiratory tract of healthy individuals. However, disruption of this commensal state can lead to pneumococcal disease (PD), including pneumonia and meningitis (2). PD primarily affects young children (3), older adults (4) and the immunocompromised (5). The annual incidence of non-invasive PD ranges between 160 – 1160 per 100.000, whereas invasive PD occurs at rates of 11 – 27 per 100.000 individuals (6). Mortality from community acquired pneumococcal pneumonia ranges from 7.3% to 13.3% and increases markedly with age, particularly in individuals older than 40 years (7, 8). Invasive PD is associated with even higher mortality, reaching up to 20.8% (9).

Given the high morbidity and mortality associated with PD, vaccination represents a very important means of prevention. Currently, there are two types of pneumococcal vaccines in clinical use, purified pneumococcal polysaccharide vaccines (PPV) and polysaccharide conjugated vaccines (PCVs) (10). PPVs are comprised solely of capsular polysaccharides (PS), whereas PCVs include a protein carrier conjugated to the capsular PS (11–13). While PPVs primarily induce T cell-independent B cell responses (14), inclusion of the protein carrier in PCV facilitates T cell help (15, 16). This T cell help, promotes B cell class switching, affinity maturation and long term memory formation following PCV, but not PPV, vaccination (17, 18). Recent studies have shown that PS-specific memory B cells are associated with reduced pneumococcal carriage rates (19). Despite these differences, both vaccines offer comparable effectiveness against invasive PD in adults, with PPV23 effectiveness estimated between 37% and 51%, compared to 47 – 65% for PCV13 (20).

The serotype coverage of PCVs has increased substantially over the past few years, with the most recent iterations in clinical development covering 31 serotypes. However, the progressive increase in valency has been associated with reduced antibody titers against individual vaccine components, a phenomenon known as immunogenicity creep (21). Although the immunological mechanisms underlying immunogenicity creep are as of yet unclear, it has been suggested that competition between antigens for limited resources in the lymph node might be a contributing factor (22).

The generation of long-lived memory B cells is thought to primarily occur in germinal centers (GCs), found in secondary lymphoid organs (SLOs), including lymph nodes (23). GCs are microanatomic sites of B cell evolution and are characterized by extensive interactions between B cells, T cells and specialized follicular dendritic cells (24). Recently, lymph node fine needle aspirations (FNA) have been used to study vaccine-induced germinal center responses following SARS-CoV-2 and influenza vaccination (25–30). These studies demonstrate long-lived GC reactions, detecting vaccine-specific GC B cells and T follicular helper cells up to 6 months post-vaccination with viral antigens (25, 27, 29, 30).

To date, GC responses in the context of multivalent vaccines and PS antigens have been scantly studied. Insight into these responses could provide a greater understanding of antigenic competition within lymph nodes and help explain the immunological basis of immunogenicity creep. In the present study, PCV13-induced GC responses and their relation to the peripheral blood response were examined. Distinct response kinetics were observed between lymph nodes and peripheral blood, with GC formation occurring only after peak expansion of antigen-specific memory B cells in circulation had occurred. Moreover, while expansion of peripheral B cells was observed across all specificities, lymph node GC B cell responses were largely confined to one or two serotypes, potentially illustrating antigenic competition during GC formation.

## Results

Five healthy young adults (median age 26 years old, range: 20 – 28, n = 2 female) with no history of pneumococcal vaccination or infection were enrolled. All participants received PCV13 at baseline. Blood and lymph node FNAs were collected at baseline and 1, 2, 3, 4, 6, 8 and 12 weeks post-vaccination (Figure 1A). Lymph node size was assessed at each visit, as well as 3, 5, 10 and 12 days post-vaccination. The median baseline lymph node cortical thickness was 2 mm, peaked at 4.6 mm on day 3 post-vaccination and remained slightly enlarged throughout follow-up (Figure 1B). Similar kinetics were observed for both the lymph node long and short axis (Supplemental Figure 1A and 1B).

**Figure 1.**
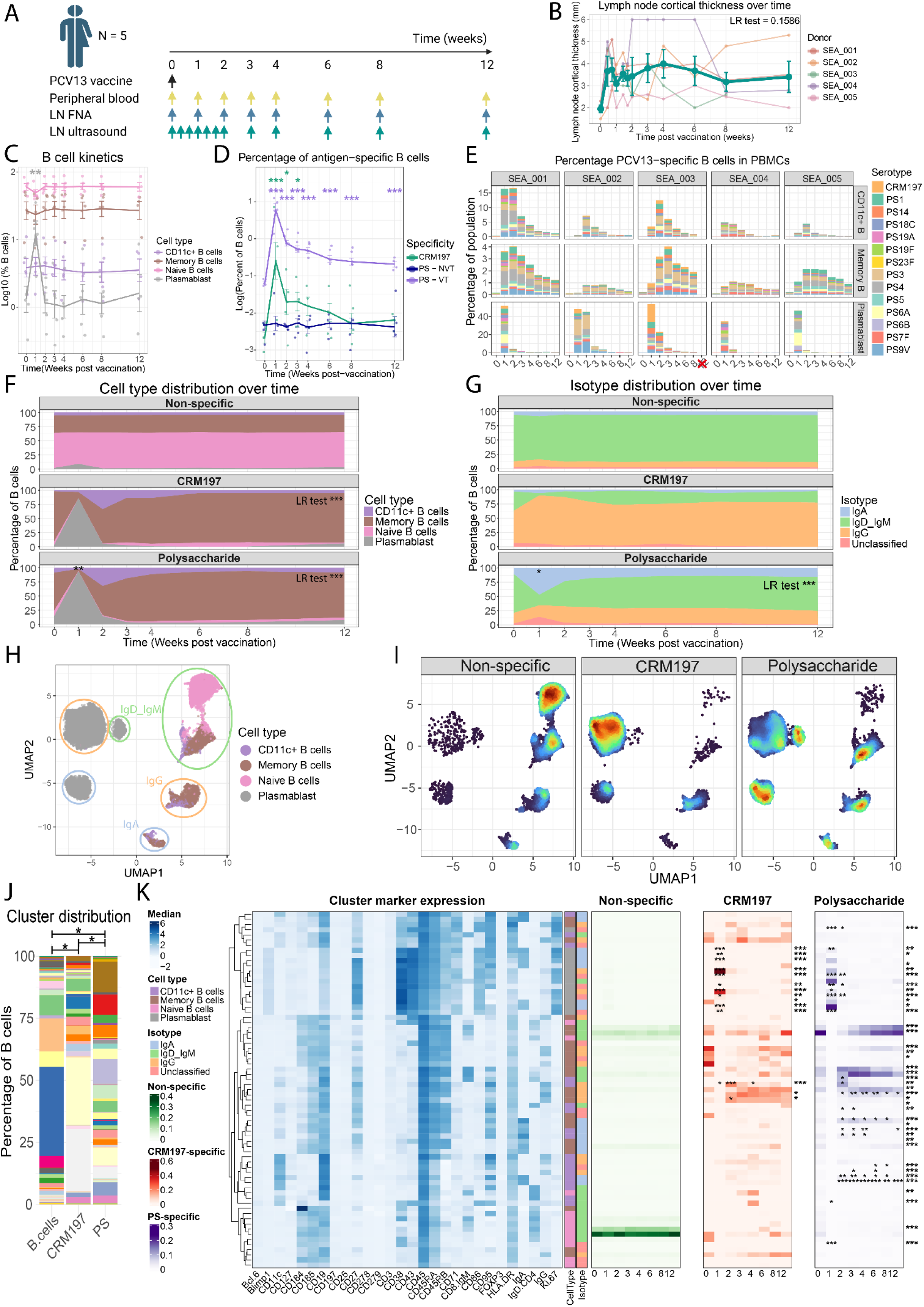
Early increase in antigen-specific B cells is observed in peripheral blood. (A) Schematic representation of the study outline and sampling time points. (B) Lymph node cortical thickness as measured by ultrasound. Bolded line represents the mean; error bars indicate standard error. (C) Main B cell subsets in peripheral blood were defined, and kinetics were plotted on a log10 scale. Data shown represents median +- SE. (D) Kinetics of antigen-specific B cells, split by antigen-specificity. Vaccine-type PS-specific B cells represents the cumulative frequency of all vaccine serotypes, whereas non-vaccine-type PS-specific B cells are PS15B-specific. Lines represent median +- SE. (E) Frequency of antigen-specific B cells are shown for all B cell subsets. Missing samples are indicated by a red cross. (F) Normalized frequencies of major B cell subsets for all specificities are plotted over time. Non-specific B cells represent B cells that are not specific for any PCV13 antigens or PS15B. Data shown represent mean frequencies across all donors. (G) Normalized frequencies of B cell isotypes are shown for each specificity over time. Unclassified indicates B cells that are negative for all of the measured isotypes. Data shown represent mean frequencies across all donors. (H) UMAP was performed of equal numbers of non-specific B cells, CRM197-specific B cells and PS-specific B cells and color-coded by phenotype. Isotype expression is highlighted by colored circles. (I) UMAP in (H), split by B cell specificity. (I) Overall cluster distribution across all time points between non-specific, CRM197-specific and PS-specific B cells Data shown represent mean across all samples. (K) Heatmap demonstrating B cell cluster abundance per specificity. Left panel shows cluster phenotype by median marker expression. Heatmaps in the right panel show normalized abundance of clusters over time, split by B cell specificity. PS = pneumococcal serotype, VT = vaccine type, NVT = non-vaccine type. Global changes were tested using a likelihood ratio test of a generalized linear mixed model (GLMM), followed by pairwise estimated marginal means (emmeans)-based comparisons upon LR test significance, comparing individual time points to baseline. Correction for multiple testing was done using the Benjamini-Hochberg method. * p < 0.05, ** p < 0.01, *** p< 0.001

### PCV13 induces short-lived plasmablast expansion in peripheral blood followed by the appearance of memory B cells

Using spectral flow cytometry, we evaluated the vaccine-induced B cell responses in peripheral blood mononuclear cells (PBMCs), measuring antigen-specificity to all 13 vaccine type PS, the carrier protein CRM197 and a non-vaccine type PS (PS15B; Supplemental Figure 1C) (31). Vaccination induced a strong but transient expansion of plasmablasts at week 1 (median frequency 1.3% at baseline vs 15% at week 1, p = 0.0021; Figure 1C). This plasmablast expansion coincided with peak antigen-specific B cell frequencies, with PS-specific B cells increasing from 0.062% at baseline to 6.1% at week 1 (p = 6.5 x 10^-14^), and CRM197-specific B cells increasing from 0.002% to 0.25% (p = 6.2 x 10^-8^, Figure 1D). Vaccine-type PS-specific B cell frequencies remained elevated up to 3 months post-vaccination (0.21% at week 12, p = 1.3 x 10^-5^). No changes were observed for the non-vaccine type PS antigen.

In order to identify which B cell subsets contributed to the antigen-specific B cell dynamics, we next examined antigen-specific B cell frequencies across B cell subsets. Antigen-specific cell frequencies, including all vaccine-type PS and CRM197, increased in all non-naïve B cell subsets 1 week post-vaccination (Figure 1E). This was most prominent in plasmablasts, which increased from cumulative 0.25% at baseline to 43% at week 1. PCV13-antigen-specific cells among CD11c^+^ B cells peaked at week 2 (median frequency 6.9%), whereas in conventional memory B cells these peaked at week 3 (median frequency 2.2%), and remained elevated up to 12 weeks post-vaccination (median frequency 0.66%). In line with these findings, PCV13-antigen-specific B cell phenotype changed dynamically over time (Figure 1F, LR test CRM197 p = 9.6 x 10^-7^, PS p = 2.3 x 10^-42^). At 1 week post-vaccination, PS-specific B cells were predominantly plasmablasts (93% of PS-specific B cells, p = 4.2 x 10^-3^), while later time points were dominated by conventional memory B cells. CD11c^+^ B cells were enriched early after vaccination (33% of PS-specific B cells at week 2 and 18% at week 3), but not later time points. Similar trends were observed for CRM197-specific responses. No temporal changes were observed for PCV13 non-specific B cells, which consisted mostly of naïve B cells (Supplemental Figure 2A).

Analysis of isotype expression revealed a transient expansion of IgA^+^ PS-specific B cells at week 1 post-vaccination, coinciding with the plasmablast expansion (% IgA^+^ 10% at baseline vs 40% in week 1, p = 0.02; Figure 2G). No temporal changes in isotype usage were observed for CRM197-specific or non-specific B cells. Overall isotype distribution differed by specificity (Supplemental Figure 2B). CRM197-specific B cells were predominantly class-switched to IgG, whereas PS-specific B cells showed a higher proportion of IgA-switched and non-class-switched memory cells. In contrast, non-specific B cells were largely non-class-switched, consistent with their predominantly naïve phenotype.

**Figure 2.**
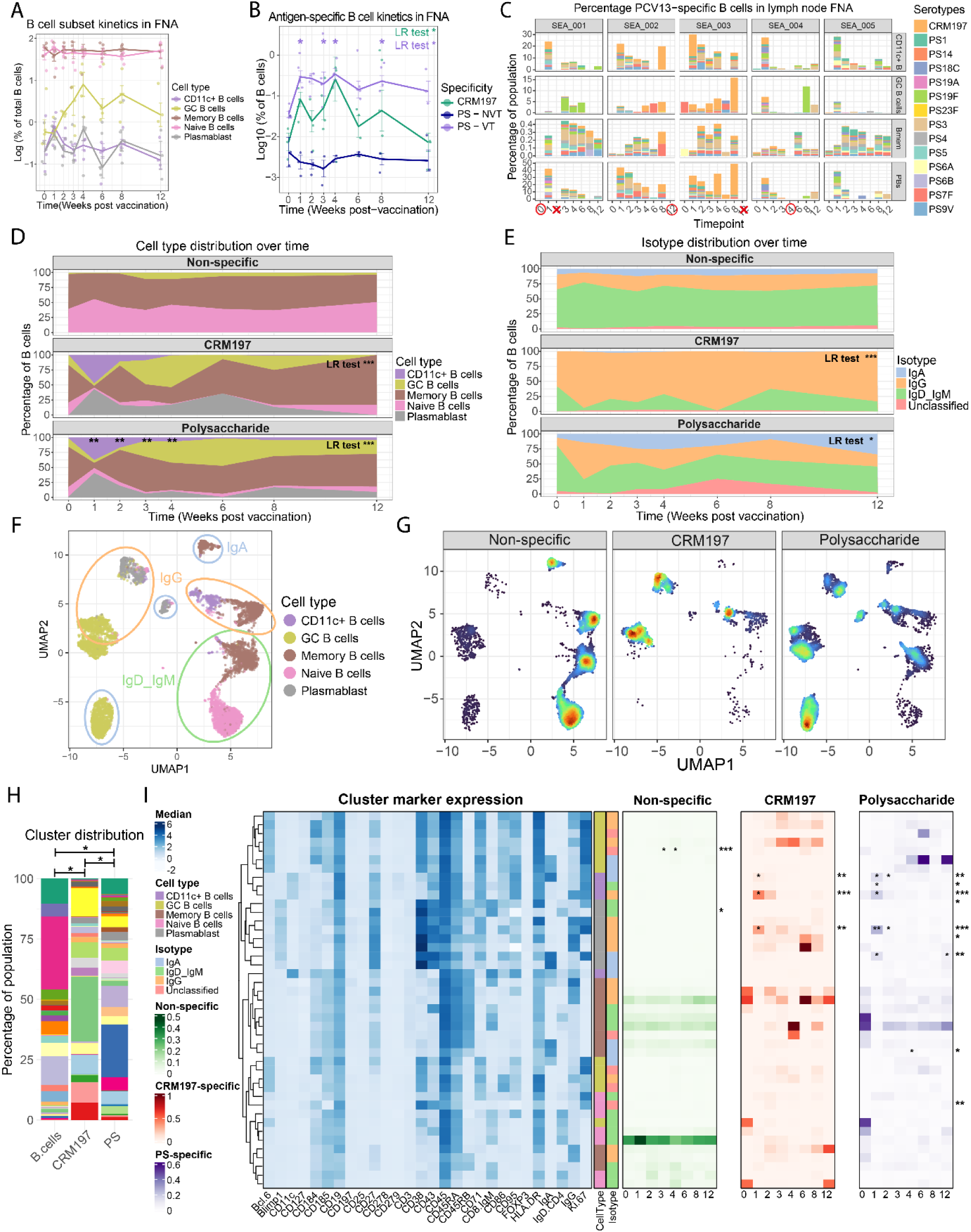
Germinal center kinetics are delayed compared to peripheral blood response. (A) Main B cell subsets in lymph nodes were defined, and kinetics were plotted on a log10 scale. Data shown represent median +- SE. (B) Kinetics of antigen-specific B cells, split by antigen-specificity. Vaccine-type PS-specific B cells represents the cumulative frequency of all vaccine serotypes, whereas non-vaccine type PS represents PS15B-specific B cells. Bolded lines represent median +- SE (C) Frequencies of antigen-specific B cells are shown for all B cell subsets. Missing samples are indicated by a red cross, while samples excluded due to high granulocyte contamination are highlighted by a red circle. (D) Normalized frequencies of major B cell subsets are plotted over time for all specificities. Non-specific B cells represent B cells that are not specific for any PCV13 antigens or PS15B. Data shown represent mean frequencies across donors. (E) Normalized frequency of B cell isotypes are shown for each specificity over time. Unclassified indicates B cells that are negative for all of the measured isotypes. Data shown represent mean frequencies across all donors. (F) UMAP was performed of equal numbers of non-specific B cells, CRM197-specific B cells and PS-specific B cells and color-coded by phenotype. Isotype expression is highlighted by colored circles. (G) UMAP in (F), split by B cell specificity. (H) Overall cluster distribution across all time points between non-specific, CRM197-specific and PS-specific B cells. Data shown represent mean across all samples. (I) Heatmap demonstrating B cell cluster abundance per specificity. Left panel shows cluster phenotype by median marker expression. Heatmaps in the right panel show normalized abundance of clusters over time, split by B cell specificity. Global changes were tested using an LR test based on a GLMM, followed by pairwise emmeans-based comparisons upon LR test significance. Correction for multiple testing was done using the Benjamini-Hochberg method. * p < 0.05, ** p < 0.01, *** p< 0.001

To integrate phenotypic and kinetic information in an unbiased manner, we next applied dimensionality-reduction and clustering approaches. Uniform Manifold Approximation and Projection (UMAP) effectively segregated B cell subsets and isotypes (Figure 1H) and revealed distinct localization of PS-specific, CRM197-specific and non-PCV13-specific B cells, highlighting their phenotypic differences (Figure 1I). Unsupervised FlowSOM clustering identified 69 B cell clusters. Cluster distribution differed significantly between specificities when combining all time points (Figure 2J), while PS-specific B cells targeting different serotypes were phenotypically more similar (Supplemental figure 2C). Analysis of B cell phenotype kinetics revealed dynamic changes over time for both CRM197-specific and PS-specific B cells (Figure 1K, Supplemental Figure 2D). Proliferating Ki67^+^ plasmablast clusters were enriched at week 1 post-vaccination for both CRM197-specific and PS-specific B cells, while several CD11c^+^ clusters increased at weeks 2 and 3 among PS-specific B cells. PS-specific memory B cell clusters expanding early after vaccination consistently expressed high levels of CD95 as opposed to CD95^-^ memory B cells at baseline and later post time points.

Overall, PCV13-vaccination induced rapid antigen-specific plasmablast expansion at week 1, followed by expansion of CD11c^+^ memory B cells at week 2 and a peak of CD27^+^ conventional memory B cells at weeks 2-3 post-vaccination. Differences between PS-specific and CRM197-specific B cells were primarily driven by isotype usage, with PS-specific B cells comprising a mixture of IgA^+^, IgG^+^ and non-class-switched memory B cells, while CRM197-specific cells were primarily IgG^+^.

### Antigen-specific B cells kinetics differ in lymph nodes compared to peripheral blood

To evaluate germinal center kinetics, we performed spectral flow cytometry on lymph node FNA samples using the same antibody panel as for PBMCs. The median FNA yield was 1.5 x 10^6^ cells (range 0.2 x 10^6^ – 8 x 10^6^ cells; Supplemental Figure 3A), with most samples passing quality control (>80% lymphocytes; 35/38 samples; Supplemental Figure 3B).

Next, we characterized the composition and temporal dynamics of lymph node B cell subsets after vaccination through manual gating of major B cell subsets (Supplemental Figure 3C). Memory B cells and naïve B cells predominated at all time points (Figure 2A). In contrast, GC B cells increased markedly, peaking at 4 weeks post-vaccination (median frequency 9.6% vs 0.58% at baseline) and gradually declining to 1.5% by week 12. Antigen-specific B cell frequencies increased over time for both vaccine-type PS- and CRM197-specific B cells (Figure 2B). PS-specific B cells rose from 0.04% at baseline to 0.23% at week 1 and remained elevated through week 8 (median frequency 0.23%), while CRM197-specific B cells increased from 0.005% at baseline to 0.023% at week 1 and persisted until week 8 (median frequency 0.04%).

To improve our understanding of the distribution of these antigen-specific responses across distinct B cell subsets, we then examined antigen-specific B cell frequencies at the subset level (Figure 2C). At week 1 post-vaccination, nearly half of all lymph node plasmablasts were antigen-specific (median antigen-specific frequency 40%), and – unlike in peripheral blood – antigen-specific plasmablasts remained detectable at later time points (median 13% at week 4). Antigen-specific CD11c^+^ B cells also expanded rapidly in the lymph node, peaking at week 1 (median frequency 29%). In contrast, PCV13-antigen-specific GC B cells increased at later time points, in accordance with total GC B cell kinetics, reaching 3.2% of GC B cells at week 4 and 4.9% at week 8 post-vaccination. Consistent with these findings, antigen-specific B cell phenotype changed over time (LR test p = 3.7 x 10^-4^ for CRM197 and p = 4.2 x 10^-6^ for PS, Figure 2D), while no changes were observed for non-specific cells. PS-specific B cells were enriched for plasmablasts and CD11c^+^ B cells at weeks 1 and 2, followed by a shift toward GC B cells from week 3 onward. Similar trends were observed for CRM197-specific B cells, albeit non-significantly. Overall, antigen-specific cells were enriched for GC B cells, CD11c^+^ memory B cells and plasmablasts compared with non-specific cells (Supplemental Figure 4A).

Assessment of B cell isotype distribution revealed significant temporal changes for both PS-specific and CRM197-specific B cells (Figure 2E, LR test p = 8.6 x 10^-5^ and 0.016 for CRM197 and PS, respectively). Both specificities showed a shift towards IgG^+^ B cells 1 week post-vaccination (CRM197: median 50% at baseline vs 95% at week 1; PS: 13% at baseline and 59% at week 1). Similar to peripheral blood, isotype usage differed by specificity (Supplemental Figure 4B). CRM197-specific were predominantly IgG^+^, while PS-specific B cells also included IgA^+^ and non-class-switched B cells.

To further resolve the phenotypic complexity of these antigen-specific B cells, we applied unbiased dimensionality-reduction and clustering approaches. UMAP analysis effectively separated major B cell subsets and isotypes (Figure 2F) and revealed distinct distributions of non-specific, PS-specific, and CRM197-specific B cells (Figure 2G). Unsupervised FlowSOM clustering identified 43 B cell clusters (Figure 2H, I), with significantly different cluster distribution across the three specificities, further highlighting their phenotypic differences (Figure 2H). Lymph node antigen-specific B cells displayed dynamic phenotypic changes over time (Figure 2I, Supplemental Figure 4C). Several CD11c^+^ memory and plasmablast clusters were enriched at week 1 post-vaccination for both CRM197- and PS-specific B cells. Notably, an IgA^+^ plasmablast cluster characterised by high CD86 and CD71 expression, along with a non-class-switched CD11c^+^ B cell cluster, were uniquely enriched among PS-specific B cells. GC B cell phenotype also differed by specificity. While CRM197-specific GC B cells were predominantly IgG^+^ with lower CD184 expression, consistent with a light zone phenotype, PS-specific GC B cells were primarily IgA^+^ and expressed higher levels of CD86, indicative of a dark zone phenotype.

In summary, vaccination induced an early expansion of antigen-specific plasmablasts and CD11c^+^ B cells within lymph nodes, followed by a delayed GC response beginning around week 3. GC activity peaked at week 4 post-vaccination and persisted for at least 2 months.

### B cell antigen-specificity differs between peripheral blood and lymph nodes

Next, we compared antigen-specificity between peripheral blood and lymph node B cells to assess whether lymph node responses mirror those observed in peripheral blood. Serotype distribution was calculated as the proportion of serotype-specific B cell of the total antigen-specific B cell pool. While peripheral blood B cells showed relatively even serotype distribution, lymph node B cells exhibited a more restricted expansion of specificities in three of five donors (Figure 3A). This reduced diversity was confirmed by significantly lower Pielou evenness indices in lymph node B cells compared with peripheral blood B cells (Figure 3B, p = 3.9 x 10^-5^).

**Figure 3.**
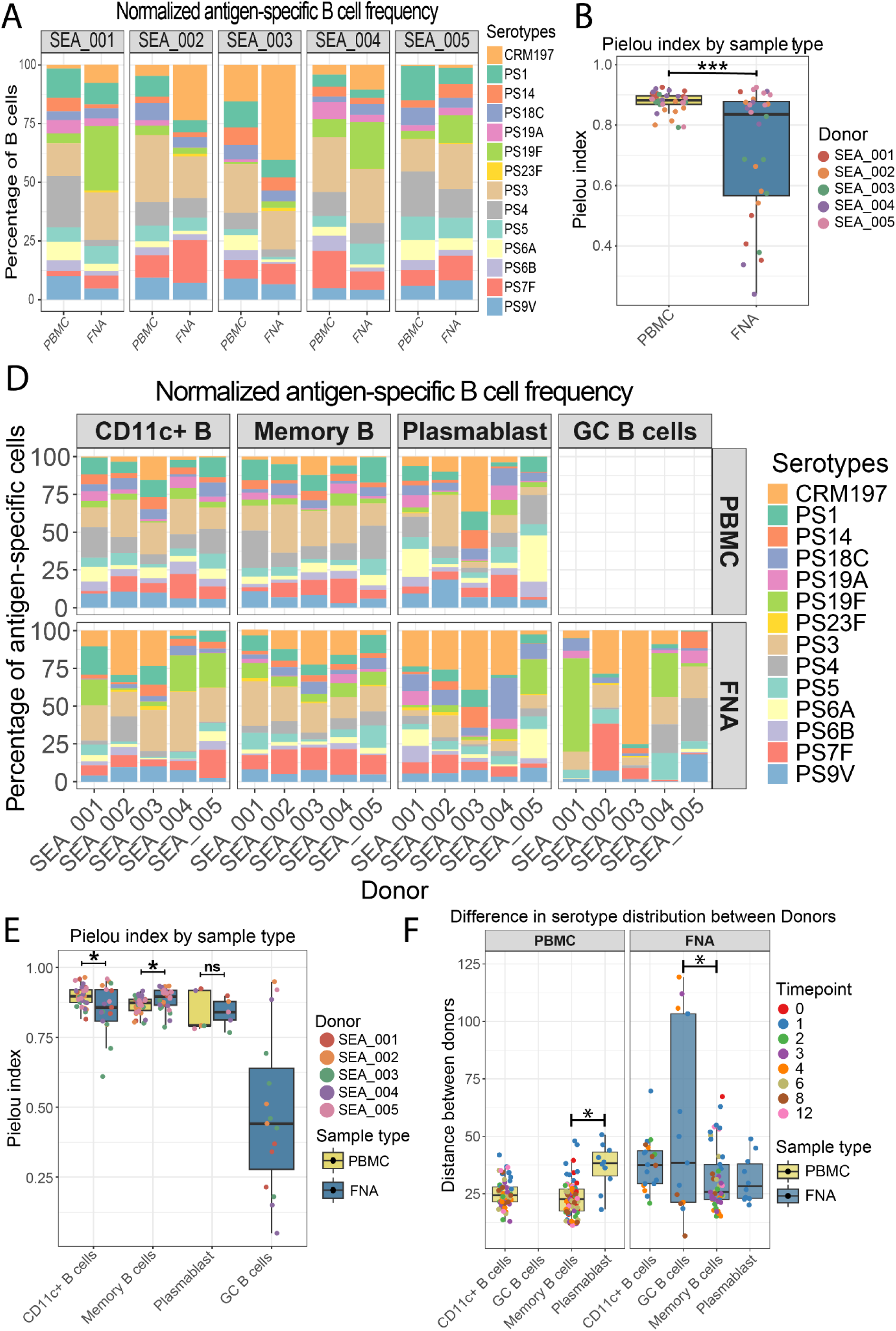
Differential specificity distribution between peripheral blood B cells and lymph node B cells. (A) Serotype distribution of antigen-specific B cells in PBMC and lymph node FNA. Data shown represent mean normalized frequency across all post-vaccination time points. (B) Pielou index for serotype distribution was calculated for PBMC and FNA samples. (C) Normalized serotype distribution of antigen-specific B cells in PBMC and lymph node FNA, split by cell type. Data shown represent mean normalized frequency across all post-vaccination time points for memory B cells, CD11c^+^ B cells and GC B cells. Plasmablast data represent serotype distribution at week 1 post-vaccination. (D) Pielou index for serotype distribution was calculated per cell type for PBMCs and FNAs. (E) Difference in serotype distribution between donors were estimated for each cell type using Euclidean distance measures in PBMCs and FNA. Differences between groups were assessed using a GLMM followed by emmeans post-hoc comparisons to compare the different groups. Correction for multiple testing was done using the Benjamini Hochberg method. * p < 0.05, ** p < 0.01, *** p< 0.001

To identify which B cell subset drove this reduced evenness in the lymph node, we examined serotype distribution within each B cell subset. Notably, GC B cell responses were dominated by a single specificity in two donors (PS19F and CRM197 for donor 1 and donor 3, respectively), and by a small number of serotypes in the other three donors (CRM197, PS2, PS3 and PS7F for donor 2; PS19F, PS3, PS4 and PS5 for donor 4; PS3, PS4 and PS9V for donor 5) (Figure 3C, Supplemental Figure 4D). In contrast, serotype distributions among CD11c^+^ B cells, memory B cells and plasmablasts were similar between blood and lymph nodes, indicating that lower total B cell numbers in FNAs were not underlying the reduced evenness. The same dominant GC specificity persisted across multiple time points in most donors, suggesting seeding by a stable GC B cell population (Supplemental Figure 4D). Consistent with this reduced evenness, GC B cells displayed a substantially lower Pielou evenness index compared to other B cell subsets in the lymph node and peripheral blood (Median Pielou index of GC B cells 0.44 compared to e.g. 0.87 for peripheral blood memory B cells; Figure 3D, Supplemental Figure 4E).

Finally, we assessed inter-donor variation in serotype distribution using Euclidean distance measures. Lymph node B cells displayed greater inter-donor variability in serotype distribution than peripheral blood B cells (Supplemental Figure 4F, p = 8.3 x 10^-13^). This increased donor variability was most noticeable for CD11c^+^ B cells and GC B cells, while memory B cells and plasmablasts had similar values between PBMCs and lymph node FNA (Figure 3E). In peripheral blood, donor variability was higher among plasmablasts compared to memory B cells (p = 2 x 10^-2^). In lymph nodes, on the other hand, GC B cells showed greater donor-to-donor variation than memory B cells (p = 2 x 10^-2^), likely reflecting their tendency to respond to a limited number of serotypes.

Taken together, these data indicate that PCV13 vaccination induces a broad and relatively uniform expansion of PS-specific memory B cells in peripheral blood across donors. In contrast, lymph node GC responses are confined to a limited number of specificities and exhibit greater inter-donor heterogeneity, highlighting compartment-specific differences in the PCV13-induced B cell responses.

### Lymph node antibodies have relatively high functional capacity compared to plasma

To assess local antibody production within lymph nodes, we measured vaccine-specific antibodies in FNA supernatants. First, we validated that FNA supernatants reflect the local lymph node microenvironment by measuring soluble factors using Olink® proteomics in a subset of FNA supernatant and plasma samples. Most analytes were present at similar or lower levels in FNA supernatant compared to plasma (e.g. IL-8, MCP-1, VEGF-A). However, 14 proteins were significantly enriched in FNA supernatant, including IL-2, CD5, CD40 and IL-1 alpha (Supplemental Figure 5A, 5B), supporting the suitability of FNA supernatants to study the local lymph node microenvironment.

Having established that FNA supernatants reflects the lymph node microenvironment, we quantified vaccine-specific antibody responses within this compartment. To control for dilution effects, antibody concentrations in FNA supernatants were normalized to albumin levels, which naturally accumulates in lymph nodes (Supplemental Figure 5C, 5D) (32). PCV13 vaccination increased IgG and IgM concentrations against all vaccine serotypes in both plasma and FNA supernatant, reaching maximum levels 2 weeks post-vaccination in both sample types (Figure 4A). In plasma, mean vaccine-type IgG increased from 0.76 µg/ml at baseline to 17.6 µg/ml 2 weeks post-vaccination, while IgM rose from 1.4 µg/ml to 14.1 µg/ml. Similar kinetics were observed in FNA supernatants, with IgG increasing from 0.024 µg/ml to 0.56 µg/ml and IgM from 0.059 µg/ml to 0.45 µg/ml over the same period. No significant changes in non-vaccine serotype antibody concentrations were observed in either compartment. Antibody kinetics were broadly similar across serotypes, although PS3-specific IgG responses were comparatively low, as expected (Supplemental Figure 5E).

**Figure 4.**
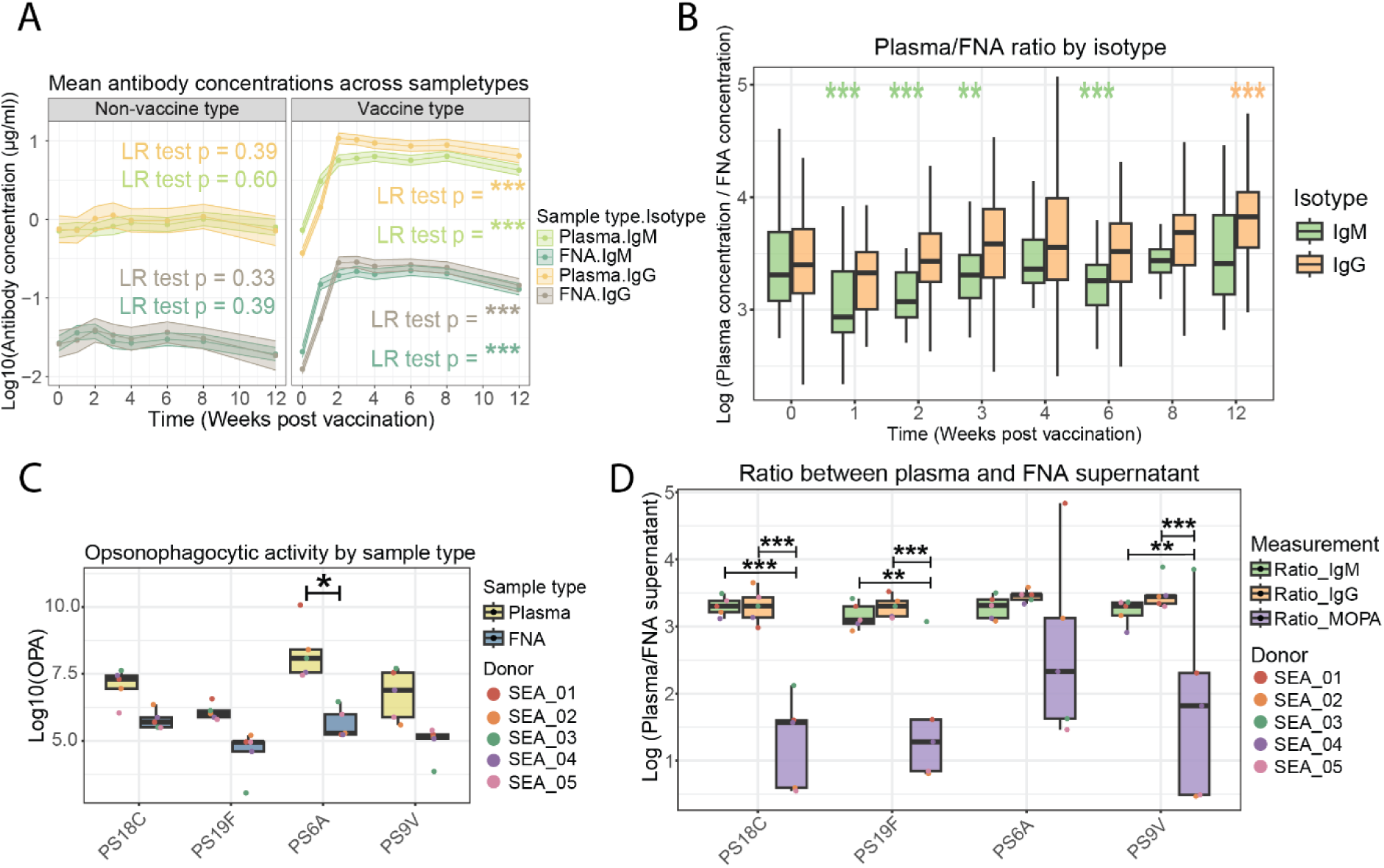
Vaccination with PCV13 induces increased PCV13-specific antibodies in peripheral blood as well as lymph nodes. (A) Mean IgM and IgG concentrations against PCV13 serotypes and non-vaccine serotypes (PS11A, PS12F, PS16F and PS33F) across plasma and FNA supernatant. Bolded lines and points show mean antibody concentration, shaded areas represent standard errors. (B) The ratio between antibody concentrations in plasma and FNA supernatant over time. Ratios were calculated separately for IgM and IgG antibodies for each donor and vaccine serotype. (E) Opsonophagocytic index for PS18C, PS19F, PS6A and PS9V in plasma and FNA supernatant. (F) Ratios between plasma and FNA supernatant were calculated for IgM and IgG concentrations as well as opsonization index and compared for each serotype. Global changes over time were assessed using a likelihood ratio test based on GLMMs. Pairwise, emmeans-based comparisons were performed upon LR test significance. Correction for multiple testing was done using the Benjamini-Hochberg method. * p < 0.05, ** p < 0.01, *** p< 0.001

To compare the temporal dynamics of antibody concentrations between compartments, where comparison of absolute concentrations may not be possible, we calculated antibody ratios between plasma and FNA supernatant (Figure 4B). For IgM, the plasma:FNA ratio decreased early post-vaccination (median ratio 3.3 at baseline vs 2.9 at week 1, p = 2.7 x 10^-10^), indicating a faster rise of antigen-specific IgM in lymph nodes than in plasma. IgM levels in FNA supernatant relative to plasma remained elevated until week 6. In contrast, IgG showed an increased plasma:FNA ratio at week 12 (ratio at baseline 3.4 vs 3.8 at week 12, p = 1.1 x 10^-4^), indicative of more rapid waning of IgG in the lymph node than in plasma.

Next, we asked whether the functional capacity of lymph node antibodies differed from those in plasma by measuring opsonization indices using multiplexed opsonization assay (MOPA) for PS6A, PS9V, PS18C and PS19F at the time of peak plasma antibody concentration. In line with antibody concentrations, opsonization indices were higher in plasma compared to FNA supernatant (Figure 4C). However, plasma:FNA ratios for opsonization indices were substantially lower than those for IgG or IgM concentrations for three out of four serotypes (Figure 4D), suggesting relatively high functional capacity of lymph node-derived antibodies compared with plasma.

In summary, FNA supernatant reflects the local lymph node microenvironment and revealed a more rapid increase in lymph node IgM levels and relatively high functional capacity of lymph node-derived antibodies.

## Discussion

Here, we describe human germinal center dynamics in the context of a multivalent polysaccharide conjugate vaccine, allowing evaluation of antigenic competition during GC formation. We show that antigen-specific B cell kinetics differ between peripheral blood and the vaccine-draining lymph node, with their expansion in peripheral blood preceding the appearance of vaccine-specific GC B cells. Moreover, antigen-specific B cells exhibited distinct serotype distributions across compartments, with GC responses in lymph nodes predominantly directed to a limited subset of serotypes.

By longitudinally sampling both vaccine-draining lymph nodes and peripheral blood at high frequency, we observed differential kinetics of antigen-specific B cells across these compartments. The expansion of peripheral blood memory B cells preceded the appearance of GC B cells, suggesting that early systemic responses are primarily driven by extrafollicular proliferation of pre-existing memory B cells. This extrafollicular response is likely to be T cell-dependent, as previous studies with PPV showed a lack of memory B cell expansion (33). Consistent with this, pneumococcal vaccination induces restricted V gene usage and comparable levels of somatic hypermutation of PS-specific B cells in vaccine-naïve individuals after vaccination with PCV and PPV, supporting a dominant contribution of pre-existing memory B cells (34). Likewise, following SARS-CoV-2 booster vaccination, memory B cells display similar levels of somatic hypermutation to those after primary immunization, further reinforcing the role of memory B cells in recall responses (35).

Early antigen-specific memory B cells frequently expressed CD11c in both peripheral blood and lymph nodes, consistent with observations after SARS-CoV-2 mRNA and influenza vaccination (36–38). Although their function remains incompletely defined, murine studies indicate that CD11c^+^ B cells are specialized for antigen uptake and presentation and localize to the T-B cell border in the spleen (39). In line with this, these cells have been shown to promote GC formation during chronic *Plasmodium* infection by priming T follicular helper cells (40). In addition, CD11c expression on B cells has been linked with an extrafollicular origin of these cells (41). The early appearance of CD11c^+^ B cells in vaccine-draining lymph nodes observed here is consistent with their extrafollicular origin and suggests a potential role for these cells in GC initiation.

Analysis of serotype distribution across peripheral blood B cell subsets revealed greater inter-donor variability in the early plasmablast response than in memory B cells and CD11c^+^ B cells. This heterogeneity likely reflects differences in pre-existing serotype-specific immunity (42–45), supported by the presence of PS-specific antibodies and memory B cells at baseline. B cell differentiation into plasmablasts has been previously shown to be affinity-dependent (46–49), and prior serotype-specific colonization may thus favor plasmablast differentiation through preferential activation of higher-affinity memory B cells. In line with this, individuals previously vaccinated with PCV exhibit enhanced plasmablast responses (50) and increased antibody production (51) upon subsequent PPV vaccination.

While we observed relatively even serotype distribution among peripheral B cell responses, lymph node GC B cell responses were largely restricted to a limited number of serotypes. In line with this, non-human primates display reduced clonal diversity in lymph nodes compared to peripheral blood following a SARS-CoV-2 booster vaccination (52).

While such clonal restriction in SLOs has traditionally been attributed to competition for limited amounts of antigen retained on follicular dendritic cells (FDCs) (53), this mechanism alone is unlikely to explain antigenic restriction after multivalent vaccination, where multiple antigens are presented simultaneously. An alternative explanation in this context may be the limited availability of GC niches within individual lymph nodes, as evidenced by murine studies showing that GCs form within pre-formed FDC clusters (54). While immunization increases FDC cluster occupancy, it does not increase their number. Accordingly, limited GC niche availability within the lymph node may contribute to antigenic restriction during multivalent vaccine responses. Together, these findings suggest that individual lymph nodes are antigenically constrained and capable of supporting GC responses to only a subset of antigens following multivalent vaccination.

Despite potential antigenic restriction within individual lymph nodes, engagement of multiple SLOs may still enable effective memory formation against multiple vaccine components. Indeed, the spleen is thought to contribute substantially to pneumococcal vaccine responses, as evidenced by reduced PS-specific B cell frequency following PCV-vaccination in splenectomized individuals (55, 56). Although clusters of lymph nodes at the vaccination site likely engage in the germinal center response (57, 58), studies suggest that GC formation in lymph nodes is anatomically restricted, showing limited GC induction in distal lymph nodes following immunization (58–60).

Together, the finite number of GC niches within SLOs (54) and the anatomic restriction of GC formation may contribute to immunogenicity creep with increasing vaccine valency (61). In this model, higher valency vaccines may saturate available GC niches, reducing the generation of affinity matured and class switched memory B cells to individual antigens. Vaccination strategies that increase recruitment of distinct SLOs may therefore help mitigate this effect. For example, contralateral boosting with a SARS-CoV-2 mRNA vaccine was shown to induce GCs that were independent of the priming site (58). Moreover co-administration of complementary pneumococcal serotypes in the contralateral limb did not impact serological response to PCV13 (62). Together, these findings suggest that spatially distributed vaccination strategies, such as bilateral administration of complementary antigens, may enhance lymph node recruitment, promote long-term memory formation and potentially overcome immunogenicity creep.

In addition to structural constraints on GC formation, local antibody availability within lymph nodes may further shape antigen retention and selection during GC responses. We observed a faster rise in vaccine-specific IgM concentrations in lymph nodes than in peripheral blood, indicating early local antibody accumulation. This likely facilitates immune complex formation and deposition on FDCs, thereby promoting GC formation (63). Consistent with this, lymph node antibodies displayed relatively high opsonizing capacity compared with plasma antibodies, potentially reflecting local production by resident antibody-secreting cells (ASCs), further enhancing antigen retention on FDCs (64–67). Differences in opsonizing capacity may in part be due explained by variations in antibody avidity between antibodies produced by lymph node resident and circulating ASCs as well as additional, unmeasured, factors within the lymph node microenvironment that enhance bacterial killing.

Despite the high sampling density and use of paired blood and lymph node samples, which enabled a detailed characterization of vaccine-induced B cell kinetics, this study has several limitations. The small sample size limits the generalizability of our findings, and variable cellular yield from lymph node FNAs led to exclusion of 3 out of 38 samples due to insufficient cell numbers. In addition, only a single vaccine-draining lymph node was sampled, and analysis of multiple lymph nodes or other SLOs could have provided a more comprehensive view of the vaccine response. Nonetheless, antigenic restriction of GC B cells was consistently observed, suggesting that this may represent a broader phenomenon. Although the low numbers of antigen-specific cells in the lymph node raise the possibility of sampling bias, samples were only analyzed when cell numbers were adequate and antigenic restriction was not observed to the same extent in other lymph node B cell subsets, suggesting this effect is limited.

Altogether, these data support a model in which recall responses to PCV13 are largely driven by extrafollicular pathways, with most antibody production and circulating memory B cells originating outside GCs. The delayed GC response likely serves to generate memory B cells that contribute to future responses upon antigen re-exposure. Finally, we observed antigenic competition in GC formation, resulting in antigenic restriction at the level of individual lymph nodes. This may provide a mechanistic basis for immunogenicity creep associated with increased vaccine valency.

## Materials and methods

### Study design and participants

In this prospective open label study, healthy, pneumococcal vaccine-naive participants between 18 and 40 years old were included. The study was performed at the Leiden University Medical Center (LUMC), Leiden, the Netherlands, between April and August 2023. All participants provided written informed consent. After obtaining informed consent, participants were vaccinated with PCV-13 (Prevenar-13) and peripheral blood and lymph node samples were collected at baseline and 1, 2, 3, 4, 6, 8 and 12 weeks post-vaccination (Figure 1A).

The study was approved by the Medical Ethical Committee Leiden – Den Haag – Delft in accordance with the EU clinical trials regulation No 536/2014 (EU CT number 2022-501519-15-00). All study procedures were performed according to the Declaration of Helsinki (amended by the 64th WMA assembly, Fortaleza, Brazil, published in JAMA 2013, volume 310, issue number 20, pages 2191 through 2194) (68) and the Medical Research Involving Human Subjects Act (WMO).

### Sample collection, preparation and storage

At each sampling time point, sodium heparinized blood was collected. Vacutainers were centrifuged for 7 minutes at 400G. Plasma was collected and stored at -70 °C following a second centrifugation for 7 minutes at 600G. Peripheral blood mononuclear cells (PBMCs) were isolated using ficoll density gradient centrifugation. Briefly, the blood pellet was diluted using Hank’s Balanced Salt Solution (HBSS; Gibco, 24020-091) and 1.077 g/mL ficoll (LUMC pharmacy) was layered underneath the diluted blood. This was centrifuged for 25 minutes at 400G, followed by collection of the interphase. Collected PBMCs were washed twice, counted manually and cryopreserved at -180 °C in 90% fetal calf serum (FCS; Serana, S-FBS-CO-015) supplemented with 10% dimethyl sulfoxide (DMSO; Merck, 1.02931.1000) at a density of 10^7^ cells per milliliter (mL).

Ultrasound-guided lymph node FNAs were performed by a qualified physician-assistant under supervision of a radiologist. The most prominent lymph node in the ipsilateral axilla was selected for FNA. Attempts were made to sample the same lymph node longitudinally using anatomical landmarks and lymph node characteristics attempts were made to sample the same lymph node longitudinally. Lymph node size and cortical thickness were measured prior to each procedure. One to two FNA passes were performed using a 21G needle and pooled into a single sample. Passes were initially collected in 500 µL Roswell Park Memorial Institute Medium (RPMI; Gibco, 42401-018) supplemented with 10% FCS, 1 mM pyruvate, 2 mM glutamate, 100 U/mL penicillin and 100 µg/mL streptomycin (R10), after which needles were flushed with 15 mL R10. FNA samples were centrifuged for 10 minutes at 400G at 4 °C, supernatant was collected, and cell pellet was pooled with the needle flushing. Following centrifugation for 10 minutes at 600G at 4 °C, FNA supernatant was stored at -70 °C. The cellular fraction was spun for 10 minutes, 400G at 4 °C, followed by erythrocyte lysis for 5 minutes at room temperature using erythrocyte lysis buffer containing 8.9 g/L of NH_4_Cl and 1 g/L KHCO_3_ (LUMC pharmacy). Cells were washed with R10, counted using Türk solution and cryopreserved in 90% FCS supplemented with 10% DMSO at a maximal density of 3 x 10^6^ cells per mL. Prior to freezing, 10% of cells were aliquoted to assess sample quality by flow cytometry as described below.

### Measurement of albumin and Olink

Soluble factors were measured in a subset of FNA supernatant and plasma samples across all donors and time points using the Olink® inflammation 96 panel. Data quality control and normalization was performed using NPX Signature software (Olink). NPX values were exported and further analyzed using Rstudio (version 2025.05.0).

Albumin concentrations in FNA supernatant were determined using the TINA-quant Albumin gen II assay (Roche, 05167043190) according to the manufacturer’s instructions. Samples were acquired using a Cobas 8000 c702 module (Roche, 06473245001).

### Measurement of antibody concentrations and opsonization activity

At the WHO Reference Laboratory for Pneumococcal Serology based at the Great Ormond Street Institute of Child Health, London, United Kingdom, plasma and FNA supernatants were assayed for IgG and IgM against vaccine-type capsular polysaccharides using a multiplexed electrochemiluminescent immunosorbent assay as previously described (69). Non-vaccine-type antibodies were measured as control, including PS12F and PS33F for both IgM and IgG as well as PS10A, PS22F and PS15A for IgM and PS6C, PS11A and PS16F for IgG.

Functional antibody capacity was measured using the Multiplexed Opsonophagocytic Assay (MOPA) (70). Briefly, heat-inactivated plasma or FNA supernatants were serially diluted and incubated with mid-log–phase bacteria in assay buffer, followed by the addition of baby rabbit complement (BRC) and differentiated HL-60 cells to facilitate opsonophagocytic killing. After incubation, phagocytosis was halted by placing the plate on ice, and each well was plated on Luria Bertani (LB) agar with 2,3,5, triphenyltetrazolium chloride (TTC) overlay. After an overnight incubation, colony forming units were enumerated using an automated colony counter, and the dilution of the sample resulting in 50% killing was calculated as the opsonic index (OI) using Opsotiter software version 3 (Bacterial Respiratory Pathogen Reference Laboratory, University of Alabama at Birmingham, USA).

### Polysaccharide and CRM197 biotinylation

Polysaccharides (PS) were biotinylated as described previously (31). All 13 vaccine serotypes as well as PS15B were included. Briefly, purified PS (SSI Diagnostica) were cyanylated using 1-cyano-4-dimethylaminopyidinium tetrafluoroborate (CDAP) and triethylamine (TAE; Sigma, T0886-100ML) to increase the pH of the solution (71). Biotinylations were performed using Pierce EZ-Link Amine-PEG_3_-Biotin (Fisher, 11881215), followed by three washes with deionized water over a 100 kDa spin filter (Merck, UFC510024) to remove unbound biotin. Biotinylated PS were resuspended in deionized water and stored at -70 °C for long term storage.

CRM197 biotinylations were performed using the EZ-Link Sulfo-NHS-LC-Biotinylation kit (ThermoFisher Scientific, 21435) according to the manufacturer’s instructions. Briefly, sulfo-NHS-LC biotin was added to purified CRM197 protein (Primrose Bio) in a 20:1 molar ratio and incubated for 2 hours on ice. Excess biotin was removed through washing three times with 400 µL deionized water using a 30 kDa spin filter (Merck, UFC503024).

Efficiency of biotin incorporation was assessed using a biotin ELISA as described previously (31). In addition, competition ELISAs were performed to confirm the maintenance of antigenicity, as described by Hoving et al (31).

### Polysaccharide and CRM197 multimer formation

Biotinylated antigens were multimerized using fluorophore-conjugated streptavidins to allow detection by flow cytometry. Combinatorial staining was used to ensure antigen specificity, as described previously (31). Fluorophore-conjugated streptavidins were added in 10 sequential steps with 10 minute intervals to obtain a final molecular ratio of 4:1. Afterwards, D-biotin (Sigma; B4501 – 100MG) was added to a final concentration of 12.5 µM to saturate streptavidin binding sites. Effective multimerization of PS was confirmed by staining beads coated with specific antisera for each of the pneumococcal PS, as described before (31). Multimers were stored at 4 °C and stored for a maximum of 2 weeks.

### Flow cytometry

All antibodies and dilutions used are listed in supplemental table 1. All staining steps were performed in the dark and Brilliant stain buffer plus (BD; 566385) was used throughout the staining procedure at the recommended concentration.

Cryopreserved cells were thawed by dropwise addition of RPMI supplemented with 20% FCS, pyruvate/glutamate and penicillin/streptomycin. Cells were washed once with thawing medium, resuspended in 200 µl phosphate buffered saline (PBS) and pipetted in 96 well V-bottom plates. Next, viability staining and CD16/CD32 blocking was performed using Live/Dead BLUE (1/1000; Invitrogen, L23105) and Fc Receptor Binding Inhibitor (1/50; eBioscience, 14-9161-73) for 15 minutes at room temperature. Cells were washed twice with fluorescence-activated cell sorting (FACS) buffer (2 mM EDTA and 0.5 % BSA in PBS) followed by incubation in 50 µl CXCR staining mix at 37 °C for 15 minutes. An equal volume of 2x concentrated cell surface staining mix was then added and incubated for another 30 minutes at room temperature. Following two washes with FACS buffer, multimer staining was performed for 30 minutes on ice. To this end, PS multimers were diluted to a final concentration of 5 µg/mL and CRM197 multimers to 0,125 µg/mL. Following two washes with FACS buffer, cells were fixed and permeabilized using the Foxp3 / Transcription Factor Staining Buffer Set (eBioscience, 00-5523-00) according to the manufacturer’s instructions. After two additional washes, cells were resuspended in intracellular staining mix, including 5x diluted multimer mix, and incubated overnight at 4 °C to improve the detection of intracellular B cell receptors (BCRs) in plasmablasts and GC B cells. The following day, cells were washed twice with perm/wash buffer and resuspended in 300 µl FACS buffer. Samples were acquired using a 5-laser Aurora spectral flow cytometer (Cytek).

### Data processing and analysis

Antibody concentrations were measured in µg/mL for both plasma and FNA supernatant and log transformed for analyses. To account for potential dilution effect and sampling variability in FNAs, normalization for protein content was performed using albumin concentration. Briefly, the correction factor for each sample was determined by calculating the ratio between the sample’s and mean albumin concentration. Raw antibody concentrations were multiplied by this correction factor to yield a corrected antibody concentration. The ratio between antibody concentrations and opsonization index in plasma and FNA was calculated to assess relative antibody concentrations.

Raw flow cytometry standard (FCS) files were unmixed using SpectroFlo software (Cytek). FNA sample quality was first assessed by determining the frequency of contaminating granulocytes among CD45^+^ cells. Samples were considered unpure and excluded from the analysis if granulocyte frequency was higher than 20%. Next, antigen-specific B cells (CD45^+^ CD3^-^ HLA-DR^+^ CD19^+^ live cells) and major B cell subsets, including CD27^+^ memory, CD11c^+^ memory, naïve, GC B cells and plasmablast were identified and gated using OMIQ (Dotmatics; see Supplemental Figure 1C and 3C for gating strategies) Unsupervised FlowSOM clustering was performed within each isotype (PBMC) or B cell subset (FNA) using Elbow metaclustering to determine the optimal number of clusters. Data, including fluorophore intensity values, antigen-specificity and cluster information, were exported and further analyzed using Rstudio (version 2025.05.0).

First, antigen cross-reactivity was determined, and cross-reactive cells were excluded from PS-specific B cell analyses. Cells were deemed cross-reactive when they were positive for more than one antigen. B cell frequencies for each specificity were determined and evaluated over time. To ensure sufficient cell numbers, phenotypic analyses were performed after pooling PS-specific B cells targeting different serotypes. Subsequently, B cell subset frequency, isotype frequency and cluster frequency were calculated within each B cell specificity and compared between non-specific cells, CRM197-specific cells and PS-specific cells. In addition, normalized serotype distribution within total antigen-specific cells were calculated and used as input for calculating the Pielou index using the Vegan R package. Using the same package, Euclidean distance measures between serotype distributions were calculated to evaluate differences in serotype distribution. To minimize the effects of sampling bias, samples were excluded when the number of antigen-specific cells was <20 for total B cell analysis or <10 for B cell subset analyses.

Generalized linear mixed models (GLMMs) using log transformed proportions were applied to evaluate changes over time, including donor ID as a random effect and time point as a fixed effect. Global changes over time or between sample types were first assessed using a likelihood ratio (LR) test, comparing the full model to a model lacking the fixed effect of interest. Upon LR test significance, post-hoc pairwise comparisons were performed using estimated marginal means (EMMs) comparisons. Changes over time were assessed relative to baseline. Correction for multiple testing was done using the Benjamini-Hochberg method.

## Data Availability

All data produced in the present study are available upon reasonable request to the authors

## Acknowledgements

We would like to extend our gratitude to the participants of the study. We thank the ultrasound technicians of the department of Radiology, LUMC, for lymph node sampling. CRM197 was kindly supplied by Primrose Bio. We acknowledge the LUMC flow cytometry core facility (https://www.lumc.nl/research/facilities/fcf) for their technical support and help with the flow cytometry analyses. This work was funded by the LUMC PhD Fellowship (to S.P. Jochems), ZonMW Vidi (to S.P. Jochems) and ZonMW Clinical Fellowship (To A.H.E. Roukens).

## Author contributions

**D.W. de Vos**: conceptualization, methodology, investigation, formal analysis, writing – original draft. **M. Johnson:** investigation. **D. Hoving:** conceptualization, methodology, writing – review and editing. **G.E. Loe-Sack-Sioe:** conceptualization, writing – review and editing. **C. Kienhuis:** formal analysis, writing – review and editing. **E.L. van Persijn van Meerten:** resources. **D. Goldblatt:** resources, writing – review and editing. **L.G. Visser:** conceptualization, supervision, writing – review and editing. **A.H.E. Roukens:** conceptualization, methodology, funding acquisition, supervision, writing – review and editing. **S.P. Jochems:** conceptualization, methodology, funding acquisition, supervision, writing – review and editing

## References

1. Ganaie F, Saad JS, McGee L, van Tonder AJ, Bentley SD, Lo SW, et al. A New Pneumococcal Capsule Type, 10D, is the 100th Serotype and Has a Large cps Fragment from an Oral Streptococcus. mBio. 2020;11(3).

2. Weiser JN, Ferreira DM, Paton JC. Streptococcus pneumoniae: transmission, colonization and invasion. Nat Rev Microbiol. 2018;16(6):355–67.

3. O’Brien KL, Wolfson LJ, Watt JP, Henkle E, Deloria-Knoll M, McCall N, et al. Burden of disease caused by Streptococcus pneumoniae in children younger than 5 years: global estimates. Lancet. 2009;374(9693):893–902.

4. Kahraman H, Yildiz P, Yilmaz S, Durmaz G, Bilgin M, Caglayan D. Impact of invasive and noninvasive pneumococcal diseases on adult populations: risk factors and vaccination status. BMC Infect Dis. 2025;25(1):172.

5. van Aalst M, Lotsch F, Spijker R, van der Meer JTM, Langendam MW, Goorhuis A, et al. Incidence of invasive pneumococcal disease in immunocompromised patients: A systematic review and meta-analysis. Travel Med Infect Dis. 2018;24:89–100.

6. Drijkoningen JJ, Rohde GG. Pneumococcal infection in adults: burden of disease. Clin Microbiol Infect. 2014;20 Suppl 5:45–51.

7. Arnold FW, Wiemken TL, Peyrani P, Ramirez JA, Brock GN, authors C. Mortality differences among hospitalized patients with community-acquired pneumonia in three world regions: results from the Community-Acquired Pneumonia Organization (CAPO) International Cohort Study. Respir Med. 2013;107(7):1101–11.

8. Ewig S, Birkner N, Strauss R, Schaefer E, Pauletzki J, Bischoff H, et al. New perspectives on community-acquired pneumonia in 388 406 patients. Results from a nationwide mandatory performance measurement programme in healthcare quality. Thorax. 2009;64(12):1062–9.

9. Chen H, Matsumoto H, Horita N, Hara Y, Kobayashi N, Kaneko T. Prognostic factors for mortality in invasive pneumococcal disease in adult: a system review and meta-analysis. Sci Rep. 2021;11(1):11865.

10. Daniels CC, Rogers PD, Shelton CM. A Review of Pneumococcal Vaccines: Current Polysaccharide Vaccine Recommendations and Future Protein Antigens. J Pediatr Pharmacol Ther. 2016;21(1):27–35.

11. Ul Haq MZ, Irfan H, Shah MSU, Sumbal A, Mughal S, Ashraf S, et al. Immunogenicity, safety and tolerability of 15-valent pneumococcal conjugate vaccine (V114) compared to 13-valent pneumococcal conjugate vaccine (PCV-13) in healthy infants: A systematic review and meta-analysis. Vaccine: X. 2025;24:100654.

12. Feng S, McLellan J, Pidduck N, Roberts N, Higgins JPT, Choi Y, et al. Immunogenicity and seroefficacy of 10-valent and 13-valent pneumococcal conjugate vaccines: a systematic review and network meta-analysis of individual participant data. eClinicalMedicine. 2023;61.

13. Pacheco-Haro M-D, de Arenas-Arroyo SN, Díaz-Goñi V, Velasco-Lucio E-J, Castellares-González C-I, Reynolds-Cortez V, et al. Immunogenicity of a 20-Valent Pneumococcal Conjugate Vaccine Versus a 13-Valent Vaccine in Infants: A Systematic Review and Meta-Analysis. Vaccines. 2025;13(11):1156.

14. Vos Q, Lees A, Wu ZQ, Snapper CM, Mond JJ. B-cell activation by T-cell-independent type 2 antigens as an integral part of the humoral immune response to pathogenic microorganisms. Immunol Rev. 2000;176:154–70.

15. Clutterbuck EA, Oh S, Hamaluba M, Westcar S, Beverley PC, Pollard AJ. Serotype-specific and age-dependent generation of pneumococcal polysaccharide-specific memory B-cell and antibody responses to immunization with a pneumococcal conjugate vaccine. Clin Vaccine Immunol. 2008;15(2):182–93.

16. Baxendale HE, Keating SM, Johnson M, Southern J, Miller E, Goldblatt D. The early kinetics of circulating pneumococcal-specific memory B cells following pneumococcal conjugate and plain polysaccharide vaccines in the elderly. Vaccine. 2010;28(30):4763–70.

17. Defrance T, Taillardet M, Genestier L. T cell-independent B cell memory. Curr Opin Immunol. 2011;23(3):330–6.

18. Obukhanych TV, Nussenzweig MC. T-independent type II immune responses generate memory B cells. J Exp Med. 2006;203(2):305–10.

19. Pennington SH, Pojar S, Mitsi E, Gritzfeld JF, Nikolaou E, Solórzano C, et al. Polysaccharide-Specific Memory B Cells Predict Protection against Experimental Human Pneumococcal Carriage. Am J Respir Crit Care Med. 2016;194(12):1523–31.

20. Farrar JL, Childs L, Ouattara M, Akhter F, Britton A, Pilishvili T, et al. Systematic Review and Meta-Analysis of the Efficacy and Effectiveness of Pneumococcal Vaccines in Adults. Pathogens. 2023;12(5).

21. Kwambana-Adams BA, Mulholland EK, Satzke C. State-of-the-art in the pneumococcal field: Proceedings of the 11(th) International Symposium on Pneumococci and Pneumococcal Diseases (ISPPD-11). Pneumonia (Nathan). 2020;12:2.

22. Dagan R, Poolman J, Siegrist C-A. Glycoconjugate vaccines and immune interference: A review. Vaccine. 2010;28(34):5513–23.

23. Liu X, Liu B, Qi H. Germinal center reaction and output: recent advances. Current Opinion in Immunology. 2023;82:102308.

24. Gatto D, Brink R. The germinal center reaction. Journal of Allergy and Clinical Immunology. 2010;126(5):898–907.

25. Turner JS, O’Halloran JA, Kalaidina E, Kim W, Schmitz AJ, Zhou JQ, et al. SARS-CoV-2 mRNA vaccines induce persistent human germinal centre responses. Nature. 2021;596(7870):109–13.

26. Scholte LLS, Leggat DJ, Cohen KW, Hoeweler L, Erwin GC, Rahaman F, et al. Ultrasound-guided lymph node fine-needle aspiration for evaluating post-vaccination germinal center responses in humans. STAR Protoc. 2023;4(4):102576.

27. Turner JS, Zhou JQ, Han J, Schmitz AJ, Rizk AA, Alsoussi WB, et al. Human germinal centres engage memory and naive B cells after influenza vaccination. Nature. 2020;586(7827):127–32.

28. McIntire KM, Meng H, Lin TH, Kim W, Moore NE, Han J, et al. Maturation of germinal center B cells after influenza virus vaccination in humans. J Exp Med. 2024;221(8).

29. Mudd PA, Minervina AA, Pogorelyy MV, Turner JS, Kim W, Kalaidina E, et al. SARS-CoV-2 mRNA vaccination elicits a robust and persistent T follicular helper cell response in humans. Cell. 2022;185(4):603–13.e15.

30. Boyd MAA, Carey Hoppe A, Kelleher AD, Munier CML. T follicular helper cell responses to SARS-CoV-2 vaccination among healthy and immunocompromised adults. Immunol Cell Biol. 2023;101(6):504–13.

31. Hoving D, Marques AHC, Huisman W, Nosoh BA, de Kroon AC, van Hengel ORJ, et al. Combinatorial multimer staining and spectral flow cytometry facilitate quantification and characterization of polysaccharide-specific B cell immunity. Commun Biol. 2023;6(1):1095.

32. Ellmerer M, Schaupp L, Brunner GA, Sendlhofer G, Wutte A, Wach P, et al. Measurement of interstitial albumin in human skeletal muscle and adipose tissue by open-flow microperfusion. American Journal of Physiology-Endocrinology and Metabolism. 2000;278(2):E352–E6.

33. Clutterbuck EA, Lazarus R, Yu LM, Bowman J, Bateman EA, Diggle L, et al. Pneumococcal conjugate and plain polysaccharide vaccines have divergent effects on antigen-specific B cells. J Infect Dis. 2012;205(9):1408–16.

34. Baxendale HE, Davis Z, White HN, Spellerberg MB, Stevenson FK, Goldblatt D. Immunogenetic analysis of the immune response to pneumococcal polysaccharide. Eur J Immunol. 2000;30(4):1214–23.

35. Li Z, Obraztsova A, Shang F, Oludada OE, Malapit J, Busch K, et al. Affinity-independent memory B cell origin of the early antibody-secreting cell response in naive individuals upon SARS-CoV-2 vaccination. Immunity. 2024;57(9):2191–201 e5.

36. Jongkees MJ, Geers D, Hensley KS, Huisman W, GeurtsvanKessel CH, Bogers S, et al. Immunogenicity of an Additional mRNA-1273 SARS-CoV-2 Vaccination in People With HIV With Hyporesponse After Primary Vaccination. The Journal of Infectious Diseases. 2022;227(5):651–62.

37. Lau D, Lan LY, Andrews SF, Henry C, Rojas KT, Neu KE, et al. Low CD21 expression defines a population of recent germinal center graduates primed for plasma cell differentiation. Sci Immunol. 2017;2(7).

38. Sutton HJ, Aye R, Idris AH, Vistein R, Nduati E, Kai O, et al. Atypical B cells are part of an alternative lineage of B cells that participates in responses to vaccination and infection in humans. Cell Reports. 2021;34(6):108684.

39. Rubtsov AV, Rubtsova K, Kappler JW, Jacobelli J, Friedman RS, Marrack P. CD11c-Expressing B Cells Are Located at the T Cell/B Cell Border in Spleen and Are Potent APCs. J Immunol. 2015;195(1):71–9.

40. Gao X, Shen Q, Roco JA, Dalton B, Frith K, Munier CML, et al. Zeb2 drives the formation of CD11c(+) atypical B cells to sustain germinal centers that control persistent infection. Sci Immunol. 2024;9(93):eadj4748.

41. Racine R, Chatterjee M, Winslow GM. CD11c expression identifies a population of extrafollicular antigen-specific splenic plasmablasts responsible for CD4 T-independent antibody responses during intracellular bacterial infection. J Immunol. 2008;181(2):1375–85.

42. Wulffraat MT, van de Weijer NG, Wijmenga-Monsuur AJ, Badoux P, Steens A, Smit D, et al. Evolving pneumococcal epidemiology and vaccine impact in the Netherlands, 2004&#x2013;2024: Carriage and invasive disease. Journal of Infection. 2025;91(6).

43. Cohen JM, Wilson R, Shah P, Baxendale HE, Brown JS. Lack of cross-protection against invasive pneumonia caused by heterologous strains following murine Streptococcus pneumoniae nasopharyngeal colonisation despite whole cell ELISAs showing significant cross-reactive IgG. Vaccine. 2013;31(19):2328–32.

44. Richards L, Ferreira DM, Miyaji EN, Andrew PW, Kadioglu A. The immunising effect of pneumococcal nasopharyngeal colonisation; protection against future colonisation and fatal invasive disease. Immunobiology. 2010;215(4):251–63.

45. Ferreira DM, Neill DR, Bangert M, Gritzfeld JF, Green N, Wright AK, et al. Controlled human infection and rechallenge with Streptococcus pneumoniae reveals the protective efficacy of carriage in healthy adults. Am J Respir Crit Care Med. 2013;187(8):855–64.

46. Phan TG, Paus D, Chan TD, Turner ML, Nutt SL, Basten A, et al. High affinity germinal center B cells are actively selected into the plasma cell compartment. J Exp Med. 2006;203(11):2419–24.

47. Wishnie AJ, Chwat-Edelstein T, Attaway M, Vuong BQ. BCR Affinity Influences T-B Interactions and B Cell Development in Secondary Lymphoid Organs. Front Immunol. 2021;12:703918.

48. Nakagawa R, Toboso-Navasa A, Schips M, Young G, Bhaw-Rosun L, Llorian-Sopena M, et al. Permissive selection followed by affinity-based proliferation of GC light zone B cells dictates cell fate and ensures clonal breadth. Proc Natl Acad Sci U S A. 2021;118(2).

49. Viant C, Weymar GHJ, Escolano A, Chen S, Hartweger H, Cipolla M, et al. Antibody Affinity Shapes the Choice between Memory and Germinal Center B Cell Fates. Cell. 2020;183(5):1298–311.e11.

50. Kättström M, Uggla B, Tina E, Kimby E, Norén T, Athlin S. Improved plasmablast response after repeated pneumococcal revaccinations following primary immunization with 13-valent pneumococcal conjugate vaccine or 23-valent pneumococcal polysaccharide vaccine in patients with chronic lymphocytic leukemia. Vaccine. 2023;41(19):3128–36.

51. Garcia Garrido HM, Vollaard A, D’Haens GR, Spuls PI, Bemelman FJ, Tanck MW, et al. Immunogenicity of the 13-Valent Pneumococcal Conjugate Vaccine (PCV13) Followed by the 23-Valent Pneumococcal Polysaccharide Vaccine (PPSV23) in Adults with and without Immunosuppressive Therapy. Vaccines. 2022;10(5):795.

52. Mesin L, Schiepers A, Ersching J, Barbulescu A, Cavazzoni CB, Angelini A, et al. Restricted Clonality and Limited Germinal Center Reentry Characterize Memory B Cell Reactivation by Boosting. Cell. 2020;180(1):92–106 e11.

53. Victora GD, Nussenzweig MC. Germinal Centers. Annual Review of Immunology. 2022;40(Volume 40, 2022):413–42.

54. Avancena P, Song T, Yao Y, Fehlner-Peach H, Diamond B, Gu H, et al. The magnitude of germinal center reactions is restricted by a fixed number of preexisting niches. Proc Natl Acad Sci U S A. 2021;118(30).

55. Gazi U, Karasartova D, Sahiner IT, Gureser AS, Tosun O, Derici MK, et al. The effect of splenectomy on the levels of PCV-13-induced memory B- and T cells. International Journal of Clinical Practice. 2018;72(5):e13077.

56. Wasserstrom H, Bussel J, Lim LC, Cunningham-Rundles C. Memory B cells and pneumococcal antibody after splenectomy. J Immunol. 2008;181(5):3684–9.

57. Barber-Axthelm IM, Kelly HG, Esterbauer R, Wragg KM, Gibbon AM, Lee WS, et al. Coformulation with Tattoo Ink for Immunological Assessment of Vaccine Immunogenicity in the Draining Lymph Node. J Immunol. 2021;207(2):735–44.

58. Burmas L, Lee WS, Kelly A, Webster R, Esterbauer R, Kent SJ, et al. Modulation of germinal center and antibody dynamics via ipsilateral versus contralateral immunization against SARS-CoV-2. The Journal of Immunology. 2025;214(3):335–46.

59. Deimel LP, Nishimura Y, Silva Santos GS, Baharani VA, Hernandez B, Oliveira TY, et al. Clonal expansion and diversification of germinal center and memory B cell responses to booster immunization in primates. Cell Reports. 2025;44(8):116142.

60. Lederer K, Bettini E, Parvathaneni K, Painter MM, Agarwal D, Lundgreen KA, et al. Germinal center responses to SARS-CoV-2 mRNA vaccines in healthy and immunocompromised individuals. Cell. 2022;185(6):1008–24.e15.

61. Hu T, Weiss T, Bencina G, Owusu-Edusei K, Petigara T. Comprehensive value assessments for new pediatric pneumococcal conjugate vaccines. J Med Econ. 2021;24(1):1083–6.

62. Simon MW, Bataille R, Caldwell NS, Gessner BD, Jodar L, Lamberth E, et al. Safety and immunogenicity of a multivalent pneumococcal conjugate vaccine given with 13-valent pneumococcal conjugate vaccine in healthy infants: A phase 2 randomized trial. Hum Vaccin Immunother. 2023;19(2):2245727.

63. Aydar Y, Sukumar S, Szakal AK, Tew JG. The influence of immune complex-bearing follicular dendritic cells on the IgM response, Ig class switching, and production of high affinity IgG. J Immunol. 2005;174(9):5358–66.

64. Oracki SA, Walker JA, Hibbs ML, Corcoran LM, Tarlinton DM. Plasma cell development and survival. Immunol Rev. 2010;237(1):140–59.

65. Tangye SG. Staying alive: regulation of plasma cell survival. Trends Immunol. 2011;32(12):595–602.

66. Kranich J, Krautler NJ. How Follicular Dendritic Cells Shape the B-Cell Antigenome. Front Immunol. 2016;7:225.

67. Krimpenfort LT, Degn SE, Heesters BA. The follicular dendritic cell: At the germinal center of autoimmunity? Cell Reports. 2024;43(3):113869.

68. World Medical Association Declaration of Helsinki: ethical principles for medical research involving human subjects. Jama. 2013;310(20):2191–4.

69. Nolan KM, Zhang Y, Antonello JM, Howlett AH, Bonhomme CJ, Greway R, et al. Enhanced antipneumococcal antibody electrochemiluminescence assay: validation and bridging to the WHO reference ELISA. Bioanalysis. 2020;12(19):1363–75.

70. Goldblatt D, Southern J, Andrews NJ, Burbidge P, Partington J, Roalfe L, et al. Pneumococcal conjugate vaccine 13 delivered as one primary and one booster dose (1 + 1) compared with two primary doses and a booster (2 + 1) in UK infants: a multicentre, parallel group randomised controlled trial. Lancet Infect Dis. 2018;18(2):171–9.

71. Zhang F, Lu YJ, Malley R. Multiple antigen-presenting system (MAPS) to induce comprehensive B- and T-cell immunity. Proc Natl Acad Sci U S A. 2013;110(33):13564–9.

