## Supplemental figures for "The germinal center B cell response to pneumococcal conjugate vaccines is antigenically restricted"

**Supplemental Table 1. Antibodies used for flow cytometry staining**

| Marker | Fluorochrome | Clone | Isotype | Vendor | Catalog # | Dilution | Stain mix |
|---|---|---|---|---|---|---|---|
| CD185 | BUV395 | RF8B2 | Rat | BD | 740266 | 25 | CXCR pre-stain |
| CD184 | BUV805 | 12G5 | Mouse IgG2a | BD | 742043 | 25 | CXCR pre-stain |
| CD25 | BV605 | BC96 | Mouse IgG1 | BD | 567571 | 50 | Surface |
| CD71 | BUV737 | LO1.1 | Mouse IgG2a | BD | 748311 | 50 | Surface |
| CD197 | BV480 | 3D12 | Rat IgG2a | BD | 566099 | 50 | Surface |
| CD127 | BB700 | hIL-7R-M21 | Mouse IgG1 | BD | 566398 | 50 | Surface |
| CD279 | RB744 | EH12.1 | Mouse IgG1 | BD | 570478 | 100 | Surface |
| CD3 | Spark Blue 574 | UCHT1 | Mouse IgG1 | Biolegend | 300488 | 100 | Surface |
| CD27 | BV785 | 0323 | Mouse IgG1 | Biolegend | 302832 | 100 | Surface |
| CD45RA | BV570 | HI100 | Mouse IgG2b | Biolegend | 304132 | 300 | Surface |
| CD38 | APC-Fire810 | HIT2 | Mouse IgG1 | Biolegend | 303550 | 300 | Surface |
| CD278 | BV750 | C398.4A | Hamster IgG | Biolegend | 313558 | 300 | Surface |
| CD95 | PE-Vio770 | DX2 | Mouse IgG1 | Miltenyi | 130-123-887 | 300 | Surface |
| CD11c | BV650 | Bu15 | Mouse IgG1 | Biolegend | 337238 | 400 | Surface |
| IgD | Pacific Blue | IA6-2 | Mouse IgG2a | Biolegend | 348224 | 200 | Surface |
| HLA-DR | BUV496 | G46-6 | Mouse IgG2a | BD | 569674 | 600 | Surface |
| CD45 | Spark YG593 | HI30 | Mouse IgG1 | Biolegend | 304084 | 600 | Surface |
| IgG | BV510 | G18-145 | Mouse IgG1 | BD | 563247 | 300 / 800 | Surface / ICS |
| CD45RB | PE | MEM55 | Mouse IgG2b | Biolegend | 310204 | 600 | Surface |
| CD86 | PE-Cy5 | IT2.2 | Mouse IgG2b | Biolegend | 305408 | 800 | Surface |
| CD8 | Spark Blue 550 | SK1 | Mouse IgG1 | Biolegend | 344760 | 1600 | Surface |
| CD19 | PE-Cy5.5 | SJ25C1 | Mouse IgG1 | eBioscience | 35-0198-42 | 800 | Surface |
| CD43 | BV421 | 1G10 | Mouse IgG1 | BD | 562916 | 1600 | Surface |
| CD4 | Pacific Blue | RPA-T4 | Mouse IgG1 | Biolegend | 300521 | 800 | Surface |
| IgM | Spark Blue 550 | MHM-88 | Mouse IgG1 | Biolegend | 314556 | 800 | Surface |
| IgA | APC-Vio770 | IS11-8E10 | Mouse IgG1 | Miltenyi | 130-113-473 | 800 / 19600 | Surface / ICS |

| | | | | | | | |
|---|---|---|---|---|---|---|---|
| FoxP3 | Spark NIR685 | 206D | Mouse IgG1 | Biolegend | 320130 | 50 | ICS |
| Bcl-6 | PE-Dazzle 594 | 7D1 | Rat IgG2a | Biolegend | 358510 | 200 | ICS |
| Blimp-1 | R718 | 6D3 | Rat IgG2a | BD | 567764 | 200 | ICS |
| Ki-67 | RB780 | B56 | Mouse IgG1 | BD | 568761 | 15000 | ICS |
| Streptavidin | BUV563 | | | BD | 612935 | | Multimer / ICS |
| Streptavidin | BV711 | | | Biolegend | 405207 | | Multimer / ICS |
| Streptavidin | BB515 | | | BD | 564453 | | Multimer / ICS |
| Streptavidin | APC | | | Biolegend | 405207 | | Multimer / ICS |
| Streptavidin | BUV615 | | | BD | 613013 | | Multimer / ICS |
| Streptavidin | BUV661 | | | BD | 612979 | | Multimer / ICS |

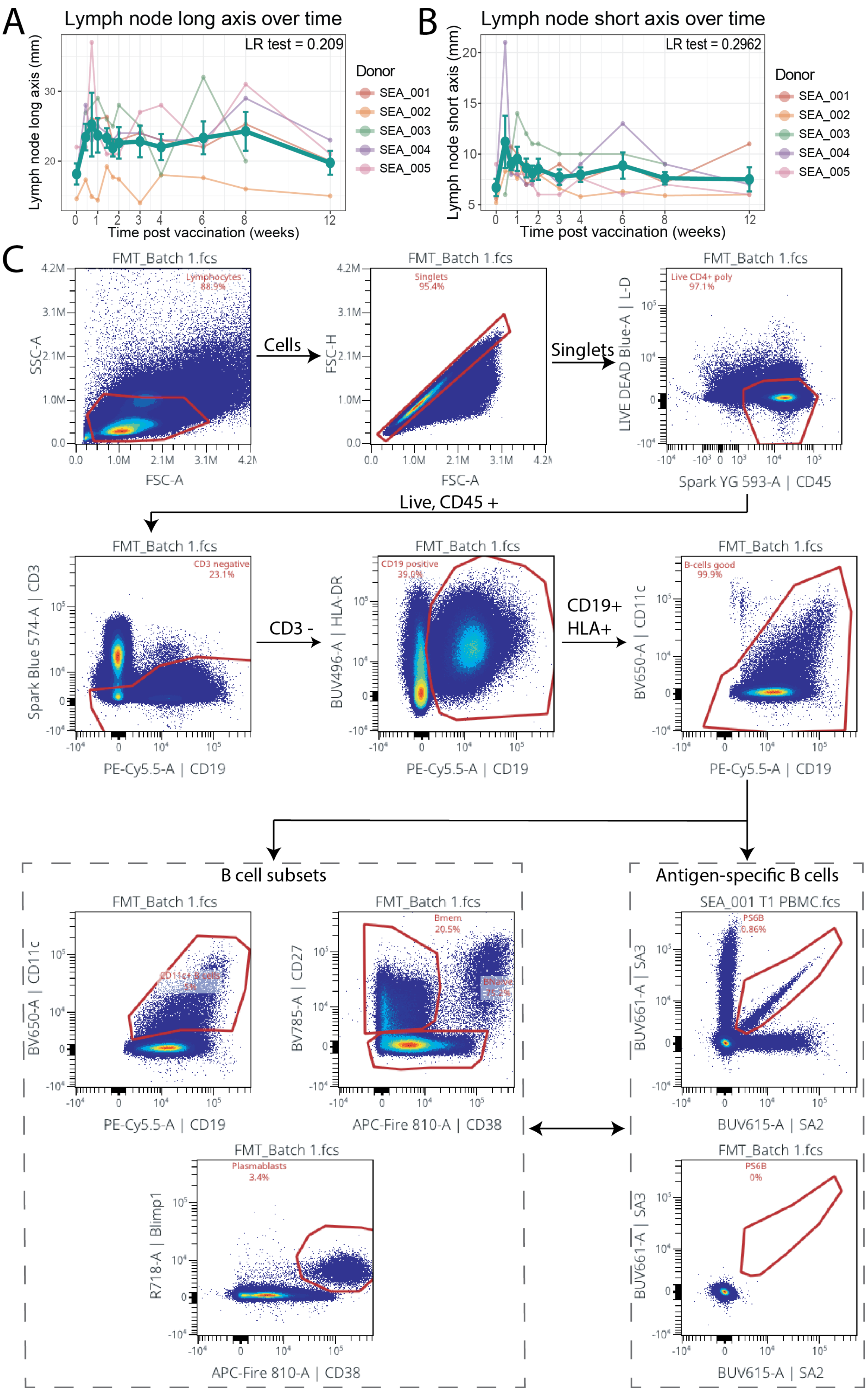
A
Lymph node long axis over time
LR test = 0.209
Lymph node long axis (mm)
Time post vaccination (weeks)
Donor
SEA_001
SEA_002
SEA_003
SEA_004
SEA_005
B
Lymph node short axis over time
LR test = 0.2962
Lymph node short axis (mm)
Time post vaccination (weeks)
C
FMT_Batch 1.fcs
Lymphocytes 88.9%
SSC-A
FSC-A
Cells
Singlets 95.4%
FSC-H
Singlets
Live CD4+ poly 97.1%
LIVE DEAD Blue-A | L-D
Spark YG 593-A | CD45
Live, CD45 +
CD3 negative 23.1%
Spark Blue 574-A | CD3
PE-Cy5.5-A | CD19
CD3 -
CD19 positive 39.0%
BUV496-A | HLA-DR
CD19+
HLA+
B-cells good 99.9%
BV650-A | CD11c
B cell subsets
Antigen-specific B cells
CD11c+ B cells
BV785-A | CD27
APC-Fire 810-A | CD38
Bmem 20.5%
SEA_001 T1 PBMC.fcs
PS6B 0.86%
BUV661-A | SA3
BUV615-A | SA2
Plasmablasts 3.4%
R718-A | Blimp1
PS6B 0%

**Supplemental Figure 1. PBMC gating strategy.** Lymph node long axis (A) and short axis (B) as measured by ultrasound. Bolded line represents the mean; error bars indicate standard error. (C) Gating strategy to define B cell subsets and antigen-specific B cells in peripheral blood. B cells were defined based on CD19 and HLA-DR expression in $CD3^{-}$, $CD45^{+}$ live lymphocytes. Contaminant myeloid cells were removed based on low expression of CD19 and high expression of CD11c. Plasmablasts were defined based on their expression of CD38 and Blimp1, whereas CD27 and CD38 were used to define memory and naïve B cells. Antigen-specific B cells were defined using a barcoding scheme; example plot is shown for one specificity including the fluorescence minus multimers control below.

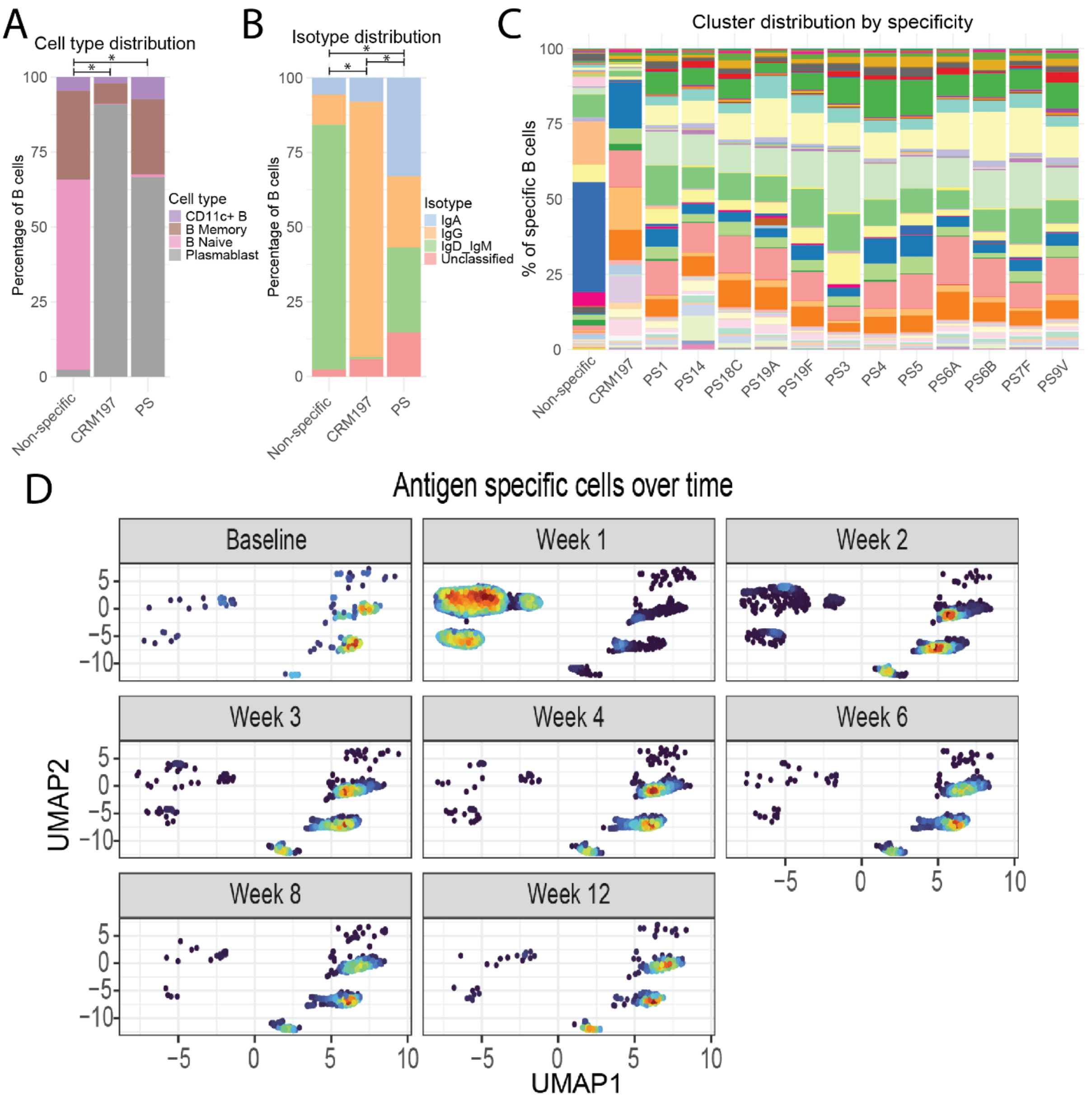


**Supplemental Figure 2. B cell phenotype in PBMCs.** (A) Major B cell subsets were defined, and overall B cell phenotype was compared between specificities. Data shown represent mean frequencies across all samples. (B) Overall isotype expression is compared between specificities. Data shown represent mean frequencies across all samples. (C) Overall cluster distribution was compared between different PS-specificities. Data shown represent mean frequencies across all time points. (D) UMAP of CRM197 and PS-specific B cells for each time point, demonstrating phenotypic changes of these cells over time.

Comparisons between groups were performed using GLMM, followed by post-hoc emmeans based comparisons. Correction for multiple testing was done using the Benjamini Hochberg method.

* $p < 0.05$, ** $p < 0.01$, *** $p < 0.001$

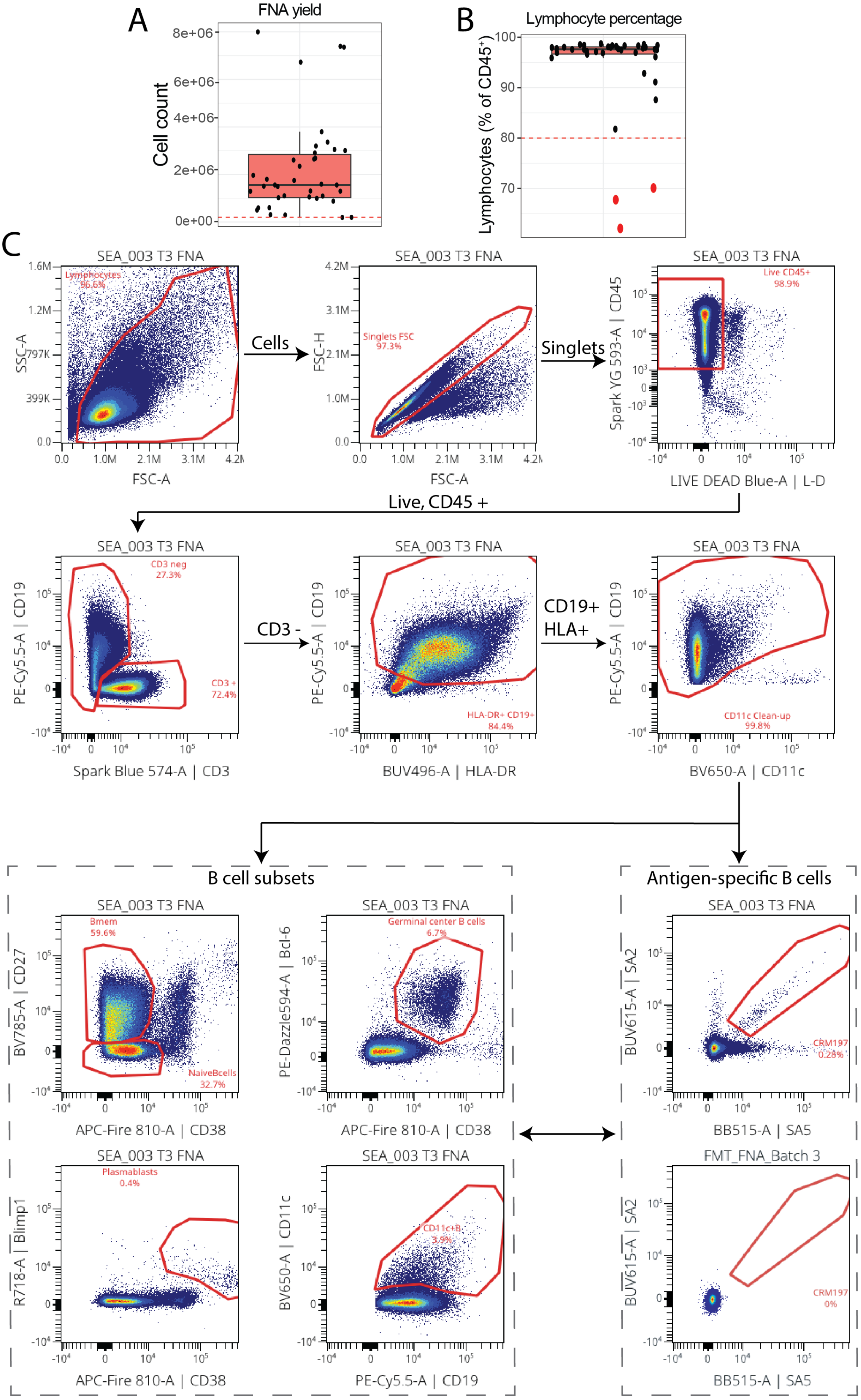
A
FNA yield
Cell count
8e+06
6e+06
4e+06
2e+06
0e+00
B
Lymphocyte percentage
Lymphocytes (% of CD45+)
100
90
80
70
C
SEA_003 T3 FNA
Lymphocytes
96.6%
SSC-A
FSC-A
Cells
Singlets FSC
97.3%
FSC-H
Singlets
Live CD45+
98.9%
Spark YG 593-A | CD45
LIVE DEAD Blue-A | L-D
Live, CD45 +
CD3 neg
27.3%
CD3 +
72.4%
PE-Cy5.5-A | CD19
Spark Blue 574-A | CD3
CD3 -
HLA-DR+ CD19+
84.4%
BUV496-A | HLA-DR
CD19+
HLA+
CD11c Clean-up
99.8%
BV650-A | CD11c
B cell subsets
Antigen-specific B cells
Bmem
59.6%
NaiveBcells
32.7%
BV785-A | CD27
APC-Fire 810-A | CD38
Germinal center B cells
6.7%
PE-Dazzle594-A | Bcl-6
CRM197
0.28%
BUV615-A | SA2
BB515-A | SA5
Plasmablasts
0.4%
R718-A | Blimp1
CD11c+B
3.9%
BV650-A | CD11c
PE-Cy5.5-A | CD19
FMT_FNA_Batch 3
CRM197
0%

**Supplemental Figure 3. Gating strategy in lymph node FNA samples.** (A) Yield of FNA samples as determined by manual counting. (B) Purity of lymph node samples was determined by fresh acquisition of FNA samples and determining the percentage of lymphocytes among $CD45^+$ events. Samples were excluded when lymphocyte percentage was < 80%. (C) Gating strategy to define B cell subsets and antigen-specific B cells in lymph node FNA. B cells were defined based on CD19 and HLA-DR expression in $CD3^-$, $CD45^+$ live lymphocytes. Contaminant myeloid cells were removed based on low expression of CD19 and high expression of CD11c. Plasmablasts were defined based on their expression of CD38 and Blimp1 and GC B cells based on CD38 and Bcl-6 expression. Memory B cells and naïve B cells were defined based on CD27 and CD38. Antigen-specific B cells were defined using a barcoding scheme; example is shown for one specificity including the fluorescence minus multimers below.

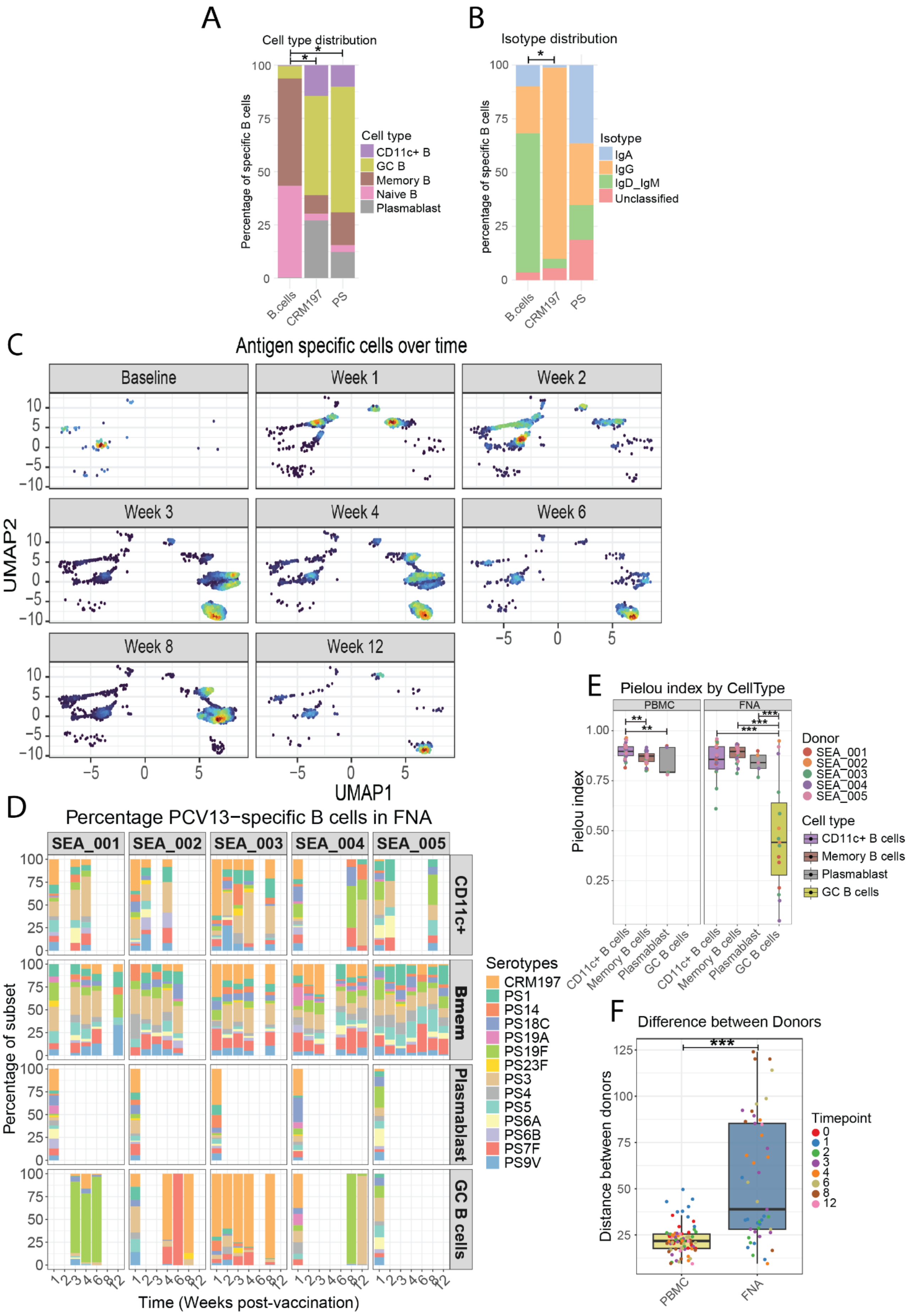

A
Cell type distribution
Percentage of specific B cells
Cell type
CD11c+ B
GC B
Memory B
Naive B
Plasmablast
B.cells
CRM197
PS
B
Isotype distribution
percentage of specific B cells
Isotype
IgA
IgG
IgD_IgM
Unclassified
C
Antigen specific cells over time
Baseline
Week 1
Week 2
Week 3
Week 4
Week 6
Week 8
Week 12
UMAP2
UMAP1
D
Percentage PCV13−specific B cells in FNA
SEA_001
SEA_002
SEA_003
SEA_004
SEA_005
CD11c+
Bmem
Plasmablast
GC B cells
Percentage of subset
Time (Weeks post-vaccination)
Serotypes
CRM197
PS1
PS14
PS18C
PS19A
PS19F
PS23F
PS3
PS4
PS5
PS6A
PS6B
PS7F
PS9V
E
Pielou index by CellType
PBMC
FNA
Pielou index
Donor
SEA_001
SEA_002
SEA_003
SEA_004
SEA_005
Cell type
CD11c+ B cells
Memory B cells
Plasmablast
GC B cells
F
Difference between Donors
Distance between donors
PBMC
FNA
Timepoint

**Supplemental Figure 4. Antigen-specific B cell phenotype in lymph node FNA samples.** (A) Major B cell subsets were defined, and overall B cell phenotype was compared between different specificities. Data shown represent mean frequencies across all samples. (B) Overall isotype expression was compared between specificities. Data shown represent mean frequencies across all samples. (C) UMAP of CRM197 and PS-specific B cells for each time point, demonstrating phenotypic changes of these cells over time. (D) Normalized serotype distribution of lymph node B cells across different B cell subsets. Subsets having less than 10 serotype-specific cells were excluded from the analysis. (E) Pielou index was compared between cell types within peripheral blood and lymph nodes. Samples with <10 serotype-specific cells were excluded from the analysis (F) Euclidean distance between serotype distribution of different donors was calculated across all B cells and compared between PBMC and lymph node B cells.

Comparisons between groups were performed using GLMM, followed by post-hoc emmeans based comparisons. Correction for multiple testing was done using the Benjamini Hochberg method.

* $p < 0.05$, ** $p < 0.01$, *** $p < 0.001$

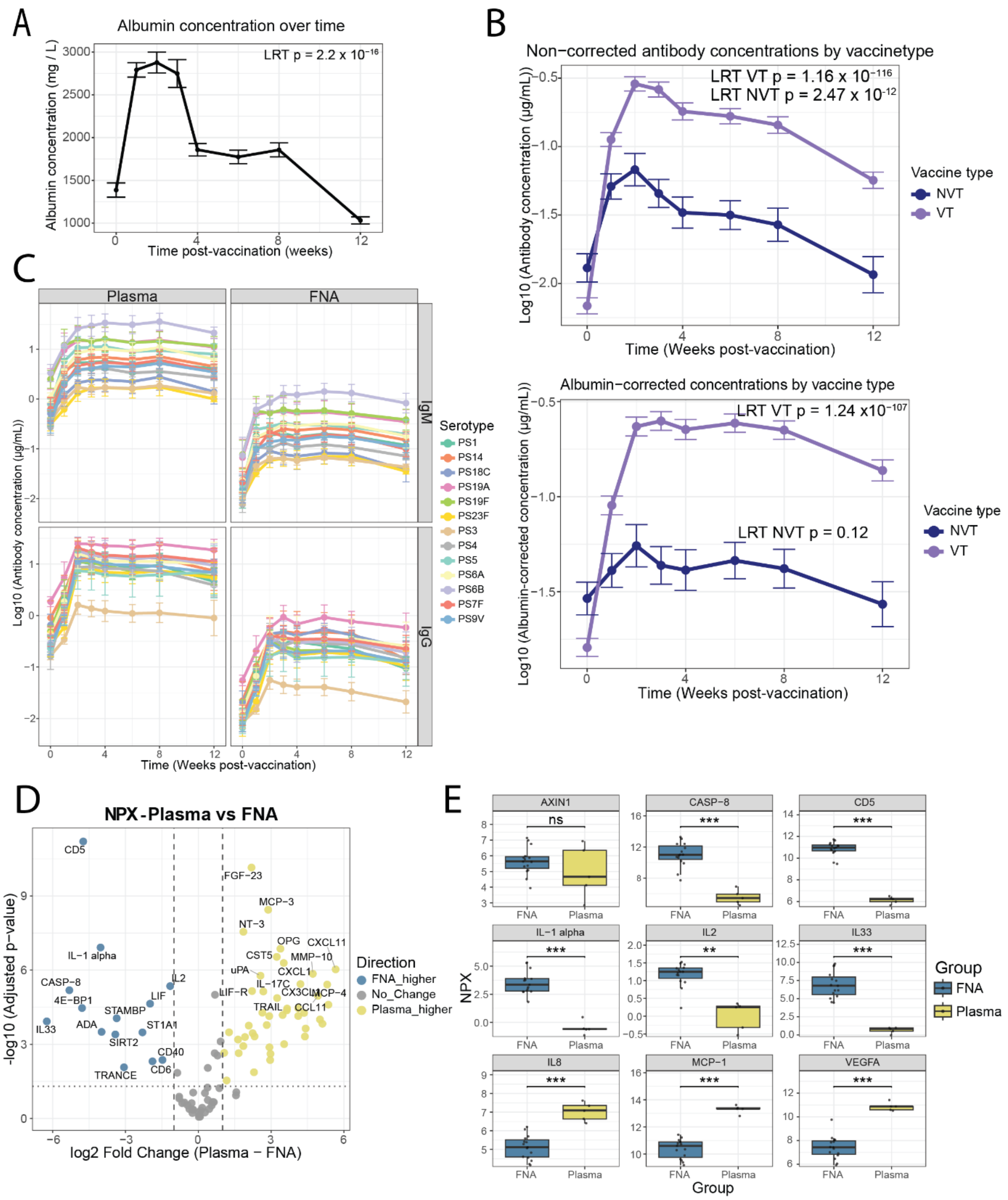


**Supplemental Figure 5. Soluble factors can be measured in FNA supernatant.** (A) Albumin concentration was measured in FNA supernatant as a measure for total protein content. Data shown represent median albumin concentration over time +- SE. (B) Antibody concentration for vaccine-type and non-vaccine-type pneumococcal serotypes were measured in lymph node FNA supernatant. Upper panel shows uncorrected antibody concentrations, whereas lower panel shows antibody concentrations after correction for albumin content. Data shown represent mean concentration across all specificities and donors +- SE. (C) IgM and IgG concentration against all vaccine serotypes as measured in plasma and FNA supernatant. Lines and points indicate mean concentration; error bars signify standard error. (D) Volcano plot comparing NPX values of soluble factors between plasma and

FNA supernatant. Colored dots represent factors that differ significantly between matrices. Top 15 hits on either side were annotated. (E) NPX values as measured by Olink, highlighting some of the top hits identified in (D).

Global time effects were tested using LR test based on GLMMs. Differences between plasma and FNA NPX were assessed using a GLMM, followed by posthoc emmeans-based comparisons. Correction for multiple testing was done using the Benjamini Hochberg method.

* $p < 0.05$, ** $p < 0.01$, *** $p < 0.001$
